# Exploring Attitudes of Primary Caregivers Towards Pediatric Tissue-Based Research using Large Language Models: Insights from Rural and Urban Community Calls and Surveys

**DOI:** 10.64898/2026.07.08.26357557

**Authors:** Anum Chotani, Nazanin Moradinasab, Brandy H. Sullivan, Lauren Griffin-Scudari, Joey Cohen, S. Fisher Rhoads, Claudia Meyer, Initha Setiady, Angela Weinhouse, Michelle Dumont, Adam R. Greene, Jay R. Thiagarajah, Jocelyn Silvester, Sarah C. Glover, Sana Syed

## Abstract

**Objectives:** Explore the perspectives of primary caregivers towards pediatric tissue-based research participation.

**Design:** Cross-sectional.

**Setting:** Two academic pediatric gastroenterology sites in the United States, including one serving a largely rural referral population and one urban clinic population.

**Participants:** Primary caregivers of children who underwent endoscopy between 2017-2018 at UVA or were seen in the clinic setting between 2024-2025 at Tulane and referred by their child’s gastroenterologist to complete an electronic survey.

**Measures and Analysis:** Primary caregiver attitudes, motivations, and concerns toward pediatric tissue-based research were explored using descriptive-focused coding in NVivo and a large language model (LLM) processing pipeline based on OpenAI’s GPT-4 for thematic, emotional, and sentiment analyses.

**Results:** Data were analyzed from 92 primary caregivers. Overall, respondents were amenable to having their children provide specimens for research. Primary motivations included a desire to help others or advance science, and perceived medical benefits for their child so long as specimen collection did not cause additional distress. Discomfort with participation was often linked to prior traumatic clinical experiences, concerns about additional biopsies causing unnecessary discomfort, or privacy issues. A desire to help others and potentially their own child was the strongest motivator for participation, while scheduling constraints and perceived risks to the child’s health were the main barriers.

**Conclusions:** At both sites, primary caregivers expressed strong willingness to participate in pediatric research. Primary concerns included perceived invasiveness of biospecimen collection and potential for additional discomfort. Limitations of the study included the unstructured nature of the data making the analysis and interpretation challenging. Strengths included two demographically diverse sites, intentional enrollment of primary caregivers of children both with and without invasive diagnostic testing, and use of LLM based analyses.

## 1. INTRODUCTION

In pediatric clinical research, primary caregivers occupy a uniquely consequential role: they oversee the health and care of their children, weigh risks and benefits on behalf of their children, and ultimately determine whether their child participates. Despite the central role caregivers play in research enrollment, especially in studies involving tissue-based procedures, their perspectives and attitudes toward pediatric clinical research remain insufficiently understood.

Understanding how caregivers perceive and experience research enrollment is important not only for practical recruitment considerations, but also for patient-centered research design. Patients and their families deserve research infrastructures that recognize and respond to their concerns and lived experiences. Pediatric clinical research may include survey-based studies, observational studies, interventional trials, and studies that collect biological samples. In this analysis, tissue-based research refers to studies that collect and analyze tissue samples, such as biopsy specimens, to examine disease features at the cellular and molecular levels. Researchers can use these samples for histologic evaluation and downstream molecular or “omics” analyses, which can help clarify disease mechanisms, biomarkers, and treatment response. In tissue-based pediatric research, where procedures involve perceived physical risk and discomfort, caregivers’ emotional and informational needs are likely to be particularly salient. Limited understanding of these perspectives may contribute to challenges in recruiting large numbers of pediatric research participants. This may potentially lead to underrepresentation of certain populations, reduced generalizability of study findings, and inequities in the distribution of downstream clinical benefits..

Prior studies have examined factors that influence caregiver willingness to enroll children in research. A 2023 psychometric study comparing attitudes of rural and urban caregivers in Mississippi identified meaningful differences in the factors shaping attitudes towards pediatric research [1]. Rural caregivers were more likely to emphasize social pressure, mistrust in researchers, and potential for direct benefits, suggesting that geography and community context may play a role in how families approach research decisions. Other studies have highlighted logistical and structural barriers that impact caregiver engagement with research opportunities. For example, recruitment strategies that involve trusted clinicians during routine clinical visits appear more effective than passive approaches such as email or social media outreach. Of note, non-White parents were less likely to engage when approached solely by website- or email-based outreach, underscoring the importance of trust and relational context in recruitment strategies [2]. Longitudinal cohort studies have also demonstrated that socioeconomic factors and time burden may influence continued participation in pediatric research, with families experiencing greater logistical strain demonstrating higher rates of attrition [3]. Finally, research on caregiver willingness to allow biological sampling in children further indicates that acceptance of invasive procedures varies based on the perceived clinical benefit to the child [4].

Together, these studies highlight the complex set of factors that influence caregiver decision-making in pediatric research participation. However, most of the existing literature has been conducted within single institutions or limited geographic settings, restricting the generalizability of findings. Additionally, prior work has focused largely on general research participation without exploring the emotional and experiential dimensions specific to tissue-based procedures. Existing studies are also often constrained by single methodological approaches. Few have directly compared caregiver attitudes across rural and urban communities using complementary qualitative and quantitative methods.

Addressing these gaps requires an analytic approach capable of systematically handling rich, semi-structured qualitative data across sites with different data collection modalities. Traditional qualitative methods, while valuable for interpretive depth, are time-intensive and difficult to standardize at scale. Recent studies have begun exploring the use of large language models (LLMs) to support qualitative research workflows. For example, one qualitative study compared investigator-led thematic analysis with analyses generated by ChatGPT-4o and Gemini using interviews from patients with cancer [5]. They found that LLMs efficiently identified structural and procedural patterns across narratives, while human researchers captured deeper emotional and contextual meanings within patient experiences. Their findings suggest that LLMs may complement, rather than replace, investigator-led qualitative analysis. Similarly, Mathis et al. demonstrated that a locally hosted open-source LLM (LLaMA-2-70B) could generate themes from clinical interview transcripts with moderate to substantial similarity to human-generated thematic analyses [6]. Importantly, their approach allowed analysis of protected health information without relying on external cloud-based models, highlighting the feasibility of secure LLM-assisted qualitative analysis in clinical research settings. Together, these studies suggest that LLMs may serve as effective tools for supporting qualitative analysis while maintaining the interpretive role of human investigators.

Because caregiver perspectives may be shaped by geographic and community context, we included two study sites serving distinct populations: UVA, which draws from a largely rural catchment area, and TU, which serves an urban and historically diverse population in New Orleans.

Here, we sought to understand how caregivers perceive and reason about research participation, particularly involving tissue-based procedures, across rural and urban communities in the United States. A deeper understanding of these perspectives serves as the foundation for a secondary goal: informing more equitable, family-centered recruitment strategies that reflect the actual concerns and values of the communities researchers seek to serve.

## 2. METHODS

We adopted a two-tiered analysis across the University of Virginia (UVA) and Tulane University (TU) to balance generalizability with depth of understanding. In the first tier, we applied a Hybrid Human-AI thematic framework (NVivo and GPT) to data from both sites. This ensured that primary caregivers’ motivations, concerns, and perceptions were identified consistently across different populations and data formats, strengthening the cross-site validity of our findings.

In the second tier, we conducted a fully automated thematic analysis using a standardized pipeline applied to both UVA and Tulane data to enable direct comparison of artificial intelligence (AI)-derived themes across sites. We then leveraged the richer UVA semi-structured interviews for additional AI modeling beyond thematic extraction, including consent grouping and emotion-, sentiment-, and semantic-level analyses. This UVA-specific extension provided deeper insight into the emotional tone and reasoning underlying primary caregivers’ decisions. Together, these tiers allowed us to characterize shared themes across sites while also revealing more nuanced cognitive and affective patterns in the UVA interviews that inform participation behavior. To empirically assess the extent of thematic similarity across sites, we used Cohen’s kappa to quantify intercoder agreement between Hybrid Human-AI and Fully Automated-AI models.

Because LLM outputs can vary with prompt framing, we incorporated a prompt-sensitivity analysis for thematic extraction. For primary analyses, we used a site-specific system prompt that incorporated contextual information about the study populations and data modalities (semi-structured interviews at UVA and structured surveys at Tulane) to support interpretation of responses within their clinical and community context. In parallel, we generated themes using a generic prompt containing only task-level instructions, enabling assessment of prompt sensitivity. Results obtained using the generic prompt were highly concordant with those produced under the site-specific prompting condition and are provided in the Supplementary Materials as a robustness analysis.

### 2.1 OVERVIEW

In this section, we describe the details of our approach at both study sites: the University of Virginia and Tulane University. This includes information on study participants, ethics and consent criteria, study design and data collection, data pre-processing, and data analysis. The overall framework is illustrated in **Figure 1**.

**Figure 1.**
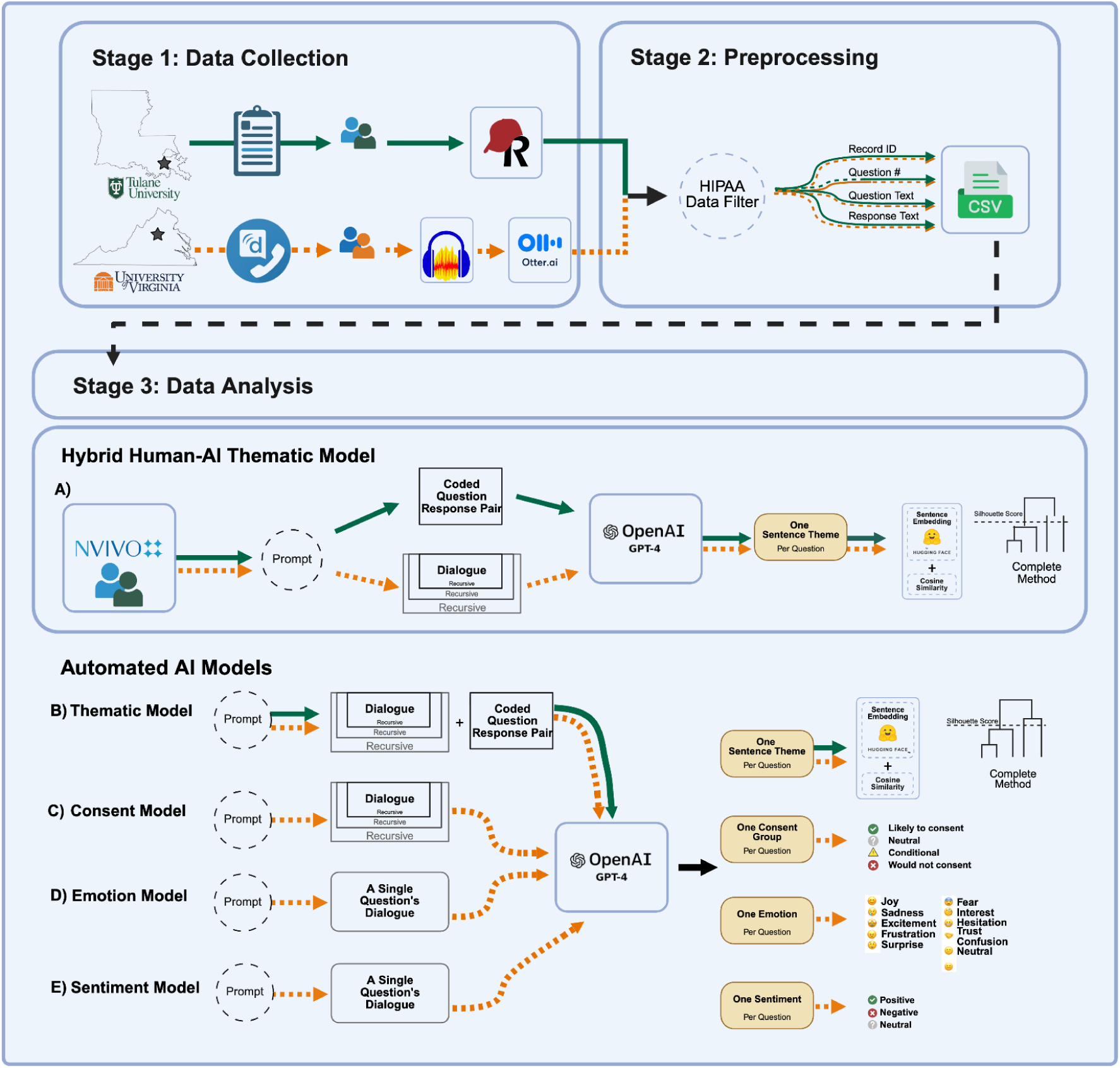
Schematic representation of study methods for Tulane University (TU; green solid line) and the University of Virginia (UVA; orange dashed line). Both UVA and TU datasets were analyzed using a Hybrid Human-AI thematic model, with UVA data undergoing additional automated GPT-4-based analyses. **Stage 1 – Data Collection:** UVA research staff conducted semi-structured telephone interviews with primary caregivers of pediatric gastroenterology patients using the Doximity Dialer and recorded audio with Audacity [7], later transcribed via Otter.ai [8] with automated speaker labeling. TU collected primary caregivers survey responses in-clinic using REDCap [23]. **Stage 2** – **Preprocessing:** Both UVA and TU datasets were manually de-identified in accordance with HIPAA guidelines. Processed records were exported as structured CSV files. **Stage 3 – Data Analysis:** Both datasets were analyzed using a Hybrid Human-AI thematic model based on NVivo and GPT-4o. In addition, both datasets underwent an Automated-AI thematic analysis using GPT-4–based large language models. Further in-depth analyses were conducted at UVA using Automated-AI GPT-4–based large language models for consent, linguistic emotion, and sentiment analysis. **(A) Thematic Model (Hybrid Human-AI):** Single-sentence themes were extracted from descriptive focused codes generated in NVivo, themes were then clustered using unsupervised hierarchical clustering with complete linkage and cosine similarity. Silhouette scoring was then utilized to determine clustering coherence. **(B) Thematic Model (Automated-AI):** Extracted a single-sentence theme per question through recursive dialogue analysis, with themes clustered using cosine similarity and hierarchical clustering (complete linkage), and cluster quality assessed via silhouette score. **(C) Consent Model (Automated-AI):** Applied recursive prompt engineering to classify responses into one of four categories—Likely to consent, Neutral, Conditional, or Would not consent. **(D) Emotional Model (Automated-AI):** Classified individual dialogue segments (without conversation history) into a single dominant emotion label. **(E) Sentiment Model (Automated-AI):** Assigned each response a single sentiment label (*positive, neutral,* or *negative*).

### 2.2 Participant Selection and Recruitment

This study was conducted at two pediatric gastroenterology sites: the University of Virginia (UVA) Medical Center in Charlottesville, Virginia, and the Manning Family Children’s Hospital affiliated with Tulane University (TU) in New Orleans, Louisiana. These sites were selected to capture caregiver perspectives across different geographic and community contexts, with UVA representing a largely rural referral catchment area and TU representing an urban pediatric gastroenterology clinic population. At UVA, the study was conducted from April 2023 to June 2024 at the UVA Medical Center in Charlottesville, Virginia. Participants were primary caregivers of pediatric gastroenterology patients. Eligible patients were identified through a retrospective chart review of electronic medical records using the Epic System. The inclusion criteria consisted of: (1) patients who underwent endoscopy between 2017 and 2018, (2) patients aged 17 years old or younger at the time of endoscopy, (3) patients who had an endoscopy showing no histopathologic abnormalities in the duodenum, (4) patients who had an endoscopy for foreign body removal, and (5) patients who had primary caregivers’ contact information available. Once eligible patients were identified, their primary caregivers were contacted by telephone and invited to participate in the study.

At TU, the study was conducted from January 2025 to April 2025 at a pediatric gastroenterology clinic within the Manning Family Children’s Hospital affiliated with Tulane University in New Orleans, Louisiana. Survey responses were collected from the primary caregivers of pediatric patients being treated at the Manning Family Children’s Hospital Gastroenterology clinic. Patients were identified through referrals from their primary gastroenterologist. Once referred, primary caregivers and primary caregivers of the patients were contacted by research staff for potential participation in the study. Inclusion criteria included caregivers of pediatric patients aged 18 years or older, who were English-speaking, and able to complete the electronic survey independently. Exclusion criteria included caregivers of pediatric patients younger than 18 years old at the time the survey was administered, non-English speaking, and limited technology literacy.

### 2.3 Ethics and Consent Criteria

At UVA, this study was approved by the Institutional Review Board of the University of Virginia (IRB # 23750). Verbal consent was obtained in lieu of written consent due to the interview taking place by telephone. At TU, this study was approved by the Institutional Review Board of Tulane University (IRB # 2023-1931). A waiver of informed consent was granted, as the survey administered electronically presented minimal risk.

### 2.4 Patient and public involvement

Patients and members of the public were not involved in the design, conduct, reporting, or dissemination planning of this study. Primary caregivers participated as study respondents by completing interviews or surveys about their attitudes toward pediatric tissue-based research.

### 2.5 Study Design and Data Collection

Data collection followed a cross-sectional study design across two academic medical centers: UVA and TU. While both sites used structured approaches to obtain caregiver perspectives, their specific data collection methods differed to align with local clinical workflows, available resources, and participant engagement procedures at each institution.

At UVA, a qualitative, cross-sectional study was conducted using semi-structured telephone interviews with primary caregivers of pediatric gastroenterology patients with no endoscopy-diagnosed small bowel disease. This process corresponds to “Stage 1: Data Collection” in **Figure 1**, where research staff conducted outreach, obtained verbal consent, and carried out interviews to gather participant responses. Five trained research staff members with clinical experience attempted to contact each identified primary caregiver up to three times. Primary caregivers who declined participation or hung up were removed from the contact list and not contacted again. Likewise, those who could not be reached after three attempts were excluded. After obtaining verbal consent, semi-structured interviews were conducted by research staff. Calls were made using the Doximity dialer to provide a UVA-specific caller ID, and the study was explained to each primary caregiver before obtaining verbal consent. Calls were made at varying times of day to accommodate varying work schedules for primary care givers. All conversations were recorded using Audacity [7] for transcription. Then, the recorded phone calls were transcribed using Otter.ai [8], which produces a text document of the conversation and assigns speaker labels for the interviewer and the participant. The interview questions were developed based on similar studies conducted by Riaz et al. [9], with additional consultation from public health experts at UVA. While a standard set of questions was asked, the semi-structured format allowed for follow-up and clarifying questions based on participant responses. The full list of interview questions is provided in **Table S.2.** In addition to the interviews, basic demographic and clinical information (e.g. patient sex, race/ethnicity, age) was extracted from each patient’s electronic medical record in **Table 1**.

**Table 1.**
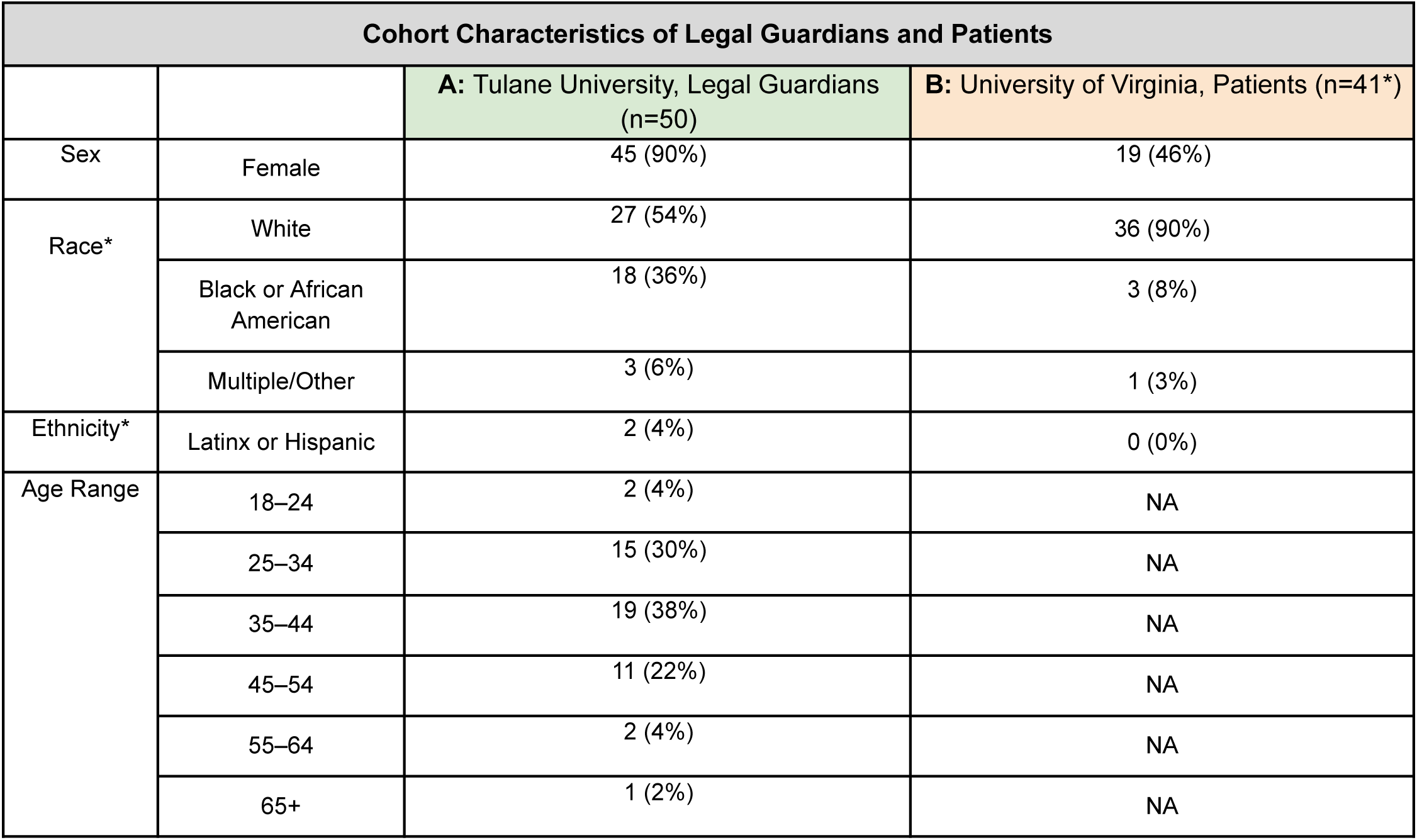
Demographic summary and cohort characteristics of legal guardians at Tulane University and patients at the University of Virginia were representative of clinical sites. **A:** Data from legal guardians that completed the survey at Tulane University. Multiple/Other races include n=1 Asian, n=1 Native American, n = 1 Unknown. **B:** Data from patients whose guardians were interviewed at the University of Virginia. *One patient at UVA did not have a self-reported race or ethnicity available in the medical record.

| Cohort Characteristics of Legal Guardians and Patients |  |  |  |
| --- | --- | --- | --- |
|  |  | A: Tulane University, Legal Guardians (n=50) | B: University of Virginia, Patients (n=41*) |
| Sex | Female | 45 (90%) | 19 (46%) |
| Race* | White | 27 (54%) | 36 (90%) |
|  | Black or African American | 18 (36%) | 3 (8%) |
|  | Multiple/Other | 3 (6%) | 1 (3%) |
| Ethnicity* | Latinx or Hispanic | 2 (4%) | 0 (0%) |
| Age Range | 18–24 | 2 (4%) | NA |
|  | 25–34 | 15 (30%) | NA |
|  | 35–44 | 19 (38%) | NA |
|  | 45–54 | 11 (22%) | NA |
|  | 55–64 | 2 (4%) | NA |
|  | 65+ | 1 (2%) | NA |

At TU, A cross-sectional study was conducted using a structured multiple-choice and open-ended survey designed by the study team to assess caregiver perspectives on childhood research participation. The survey was developed using data from studies previously conducted by Riaz, et al. [9] and research staff at the UVA, with adaptations by a sociologist and research staff at TU. The survey questions were formatted using 3- to 5-point Likert scales, with two open-ended questions. Surveys were administered in-person, electronically, via tablet devices to primary caregivers of pediatric gastroenterology patients during routine clinic visits. Two trained research staff members attended routine clinic visits once weekly to discuss the research study and obtain survey responses from primary caregivers of patients who were amenable to study participation. All survey responses were collected electronically via tablet device from each participating caregiver. Study data were collected and managed using REDCap [23] electronic data capture tools hosted at Tulane University. A complete list of survey questions is provided in **Table S.3.** In addition to the surveys, basic demographic and clinical information (e.g., the caregiver’s gender at birth, race/ethnicity, and highest level of education) was collected from the caregivers of each patient enrolled in **Table 1** and **Table S.1**.

### 2.6 Data Pre-processing

Data pre-processing followed a structured workflow to ensure accuracy, completeness, and the removal of identifiable information prior to analysis (**Figure 1**, Stage 2). All data from both study sites underwent manual quality checks and de-identification procedures.

At UVA, one research staff member manually reviewed the transcribed phone interviews to correct transcription errors and ensure that no placeholder or filler words were introduced for a total of 11 questions. The resulting text was then de-identified by removing private health information, including patient names and other directly identifying details.

At TU, electronically collected survey responses for a total of 22 questions were obtained via the REDCap database [23]. To ensure data quality, a research staff member manually reviewed each submission to verify that all questions were answered appropriately. The data was subsequently de-identified by removing protected health information such as patient and caregiver names, addresses, zip codes, and medical record numbers.

### 2.7 Hybrid Human–AI Thematic Model

We employed a Hybrid Human-AI thematic analysis approach using a human coder to extract descriptive codes from participant responses at both sites. A LLM was then utilized to generate thematic outputs and conduct additional clustering analysis. This hybrid approach was utilized to maintain accurate representation of caregiver responses across sites. This hybrid approach also mitigates the impact of overgeneralization, loss of contextual awareness, and external validity that may be compromised with a fully automated approach **(Figure 1)**.

The same analytic framework was applied after initial manual coding by a single human coder using descriptive-focused coding at both sites. Qualitative responses were first exported (UVA interview transcripts and Tulane REDCap survey outputs) and manually coded using descriptive-focused coding in NVivo using a single clinical researcher. Subsequently, a set of 24 codes for Tulane REDCap survey outputs and 41 codes for UVA OtterAI interview transcript outputs were generated (**Table S.8 and S.9, respectively**).

For TU multiple-choice questions, each predefined response (e.g., risk to child’s health, privacy concerns) was mapped to an equivalent descriptive-focused code (e.g., potential harm to child’s well-being, privacy and data security concerns). Free-text responses from both sites were also mapped to an equivalent descriptive-focused code. These NVivo-generated codes [18] were then fed into the GPT-4 omni [10] model to automatically generate single-sentence thematic summaries for each framed and standard open-ended question. GPT-4 omni outputs were subsequently grouped using unsupervised clustering and semantic-similarity analysis, producing higher-level thematic clusters. Additional hyperparameter settings and methodological details are provided in **Table S.4** and **Supplementary section B.1**.

To assess the influence of prompt specificity on theme generation, we implemented a two-system prompt configuration. A site-specific prompt framed the model as a specialized qualitative researcher and incorporated contextual guidance relevant to the UVA and TU caregiver populations, including study setting and data modality. A generic prompt provided only task-level instructions without population-specific detail, supporting cross-site comparability and minimizing context leakage. Both prompts imposed identical output constraints (i.e., one-sentence thematic summaries), and all downstream processing steps were held constant. The site-specific and generic prompts used for thematic extraction are provided in the Supplementary Materials, including the UVA site-specific prompt (**Supplementary section A.1**), UVA generic prompt (**Supplementary section A.2**), Tulane site-specific prompt (**Supplementary section A.3**), and Tulane generic prompt (**Supplementary section A.4**).

### 2.8 Fully Automated-AI Analysis

Given the conversational and highly open-ended structure of the UVA interview data, relying solely on manual or hybrid coding introduced risks of coder variability and potential loss of nuanced meaning. To address these challenges and to ensure analytic consistency across diverse narrative responses, we incorporated a suite of fully automated AI-based analyses. These methods were designed to enhance interpretability, reduce subjectivity, and uncover deeper linguistic, thematic, and affective patterns that are often difficult to capture through manual review alone.

Using GPT-4o–based large language models, we implemented automated analytic pipelines. First, we applied an Automated Thematic Model to both the UVA and Tulane datasets to derive single-sentence themes from free-text responses; themes were then clustered using cosine similarity and hierarchical clustering (complete linkage). Second, because the UVA interviews provided richer, multi-turn dialogue suitable for deeper modeling, we conducted three additional automated analyses on the UVA dataset only: (1) a Consent Model to classify responses into one of four categories—Likely to consent, Neutral, Conditional, or Would not consent—using recursive prompt engineering; (2) an Emotional Model to classify individual dialogue segments (without conversation history) into a single dominant emotion label; and (3) a Sentiment Model to assign each response a single sentiment label (positive, neutral, or negative). Each model was developed using prompt engineering and optimized settings to maintain consistency across the diverse response styles present in the UVA interviews.

#### 2.8.1 Automated-AI Thematic Model

To generate concise, high-level themes from the conversational UVA interview data and TU survey data, we developed an AI-driven thematic analysis pipeline using GPT-4–selected for its state-of-the-art language understanding, flexibility in prompt engineering, and parameter tuning. enabled us to tailor the model to our specific task, as illustrated in Stage 3 of **Figure 1**. The objective of this model was to produce a single-sentence theme for each interviewer question based solely on the participant’s most recent response, while incorporating prior conversation history only when explicitly referenced. This ensured that each theme reflected the respondent’s immediate perspective without unintended influence from earlier dialogue.

The thematic model operated using a structured prompting strategy (**Figure S.1**), in which the system prompt positioned the model as an expert qualitative researcher with experience in community-engaged research, mixed-methods analysis, pediatric caregiver contexts, and health literacy considerations. To assess the influence of prompt specificity on theme generation, we implemented a two-system prompt configuration as described in **Section 2.7**.

The user prompt provided the interview conversation history and the most recent question–response pair, and the assistant prompt produced a synthesized one-sentence theme that captured the essence of the participant’s statement.

Interview data were processed iteratively: each participant response was fed into the model along with its associated prompt framework, generating one theme per question across the entire transcript. Full hyperparameter settings, and prompt templates used in the thematic analysis pipeline are documented in the **Supplementary section B.2.**

Because one-sentence themes cannot be easily summarized across phone interviews using traditional statistical methods we conducted post-processing to identify overarching patterns. Specifically, we applied unsupervised clustering to group the generated themes for each question across all interviews. To achieve this, we used the SentenceTransformer [11] library, which includes state-of-the-art pretrained models that convert sentences into embeddings capturing semantic relationships. This allowed us to cluster similar themes—even when phrased differently—into meaningful groups. We selected the SBERT [12] framework, a version of BERT [13] optimized for sentence-level semantic understanding, making it well-suited to our task. We chose the all-MiniLM-L12-v2 [14] model specifically due to its ability to effectively capture semantic relationships while maintaining consistent clustering performance across responses. We then clustered these embeddings to group similar themes into meaningful categories, regardless of phrasing. To measure similarity between embeddings, we used cosine distance as the metric. Hierarchical clustering was applied due to its flexibility in handling an unknown number of clusters, and the optimal cluster count was determined using silhouette scores. Detailed equations, model settings, and evaluation procedures are provided in the **Supplementary section C**.

#### 2.8.2 Automated-AI Consent Model

To analyze participants’ consent behavior for each interview question and to understand their overarching stance on the interview topics, we categorized responses into four consent groups: (1) likely to consent, (2) neutral, (3) conditional, and (4) would not consent. We developed an Automated-AI Consent model using GPT-4 [10] The model takes each interview question along with the corresponding participant response and outputs a classification into the appropriate consent group. To ensure high-quality and consistent outputs, we performed careful customization of the model through hyperparameter tuning and prompt engineering. Additional details on the hyperparameter tuning are provided in **the Supplementary section D**.

The system prompt provided high-level instructions, defined the consent categories, and outlined the decision criteria used during classification. The user prompt contained the full conversation history up to the point of the response being evaluated. Including the conversation history was essential, as some responses explicitly referenced prior statements (e.g., “same as before”), which could not be interpreted correctly without earlier dialogue. The assistant prompt specified the required output format;namely, a single consent-group label for the response (e.g., “Likely to consent”). An illustrative example of the input–output structure used for the second interview question is shown in **Figure S.2.**

#### 2.8.3 Automated-AI Emotion Model

We adopted a similar preprocessing pipeline as used in the Consent and Thematic Models. The interview data was formatted as a grouped question–response pairs per transcript. However, for this Automated-AI Emotion Model, we chose to exclude conversation history when feeding data into the model. This decision was made to ensure the model’s output reflected the localized emotional tone of each response, rather than being influenced by previous exchanges within the same conversation. The parameters used for the linguistics emotion analysis match exactly to consent categorization parameters due to their ability to classify well. The prompts, however, were tailored specifically for the emotion classification task. In the system prompt, we provided contextual information about both the nature of the task and the type of data the model would receive. Since medical conversations often involve more sensitive language than conventional topics, we explicitly informed the model that medical terminology may be present. This ensured that the model was appropriately primed for interpreting the emotional tone within a clinical research interview context. The system prompt is shown in **Figure S.3**.

We selected 11 emotion categories:Joy, Sadness, Excitement, Frustration, Surprise, Fear, Interest, Hesitation, Trust, Confusion, and Neutral. Categories were chosen because they reflect a broad range of emotional expressions that participants might convey during a phone interview, particularly in the context of discussing perceptions of clinical research. The user prompt consisted of the interview dialogue corresponding to a specific question–response exchange. An example from the first transcript is shown in **Figure S.3.**

#### 2.8.4 Automated-AI Sentiment Model

The Automated-AI Sentiment Model followed the same structure as the Emotion Model, with the primary difference being the output categories. Instead of labeling specific emotions, this analysis classified each participant’s response to a given question as expressing either positive, negative, or neutral sentiment. To determine the most appropriate model for this task, we evaluated several large language models, including the IBM Watson Natural Language Understanding [15] platform, Facebook’s RoBERTa [16] model fine-tuned for sentiment analysis on Twitter data (twitter-roberta-base-sentiment) [17], and OpenAI’s GPT-4 [10] model. Consistent with our earlier analysis, we ultimately chose GPT-4 because its prompt engineering capabilities enable extensive customization. In contrast, IBM Watson [15] and twitter-roberta-base-sentiment [17] rely on a fixed set of sentiment categories, which limits their adaptability to the unique nuances of phone interview-style data. This flexibility made GPT-4 [10] the most suitable option for tailoring responses to our domain-specific needs. The model parameters remained consistent with those used in the emotion analysis (e.g., temperature, max tokens, frequency penalty, and presence penalty).

The system prompt was slightly modified to reflect the new task of sentiment classification, targeting the broader polarity categories: positive, negative, and neutral. shown in **Figure S.4.** Similarly, the user and assistant prompts remained consistent with those used in the emotion analysis, except that the assistant prompt reflected a particular sentiment rather than an emotion.

## 3. RESULTS

### 3.1. Analysis of Participant Demographics and Clinical Data

Across both study sites, UVA and TU, demographic and clinical characteristics were summarized to contextualize the populations represented in the primary caregivers’ interviews and surveys. At the UVA, a total of 42 primary caregivers were interviewed between April 2023 and June 2024. A demographic summary of the patients whose primary caregivers were interviewed is presented in **Table 1**. Slightly more than half of the patients were male (54%), and 46% were female. The majority of patients self-identified as White (90%), followed by African American (8%), and Other racial backgrounds (3%). All patients were self-reported as Non-Hispanic. Geographic distribution based on the Rural-Urban Commuting Area (RUCA) codes showed that most patients resided in metropolitan areas, with 44% living in a metropolitan core area and 32% in high-commuting metropolitan zones. A smaller proportion of patients came from micropolitan areas (12%) or more rural locations, including small towns and isolated areas (combined 5%). In terms of the clinical indications for endoscopy, the most common presenting symptom was abdominal pain (54%), followed by reflux or regurgitation (15%), and poor weight gain (12%). Less frequently reported symptoms included dysphagia (10%), diarrhea (7%), feeding difficulties (5%), and vomiting (2%).

At TU, a total of 50 primary caregivers were surveyed between January 2024 and April 2025. A demographic summary of primary caregivers who completed the survey is presented in **Table S.1**. The majority of primary caregivers were female (90%), with males comprising the 10% of participants. Over half identified as White (54%), followed by African American (8%), Hispanic or Latinx (4%), Asian American (2%), and Native American (2%). An additional 2% of primary caregivers identified as another racial or ethnic background. More than half of the primary caregivers were married (52%). The highest education level attained was a doctorate degree (8%), twice as many received a master’s degree (16%), 20% received a bachelor’s degree, and 24% received a high school diploma. Most primary caregivers (70%) had more than one child. In terms of employment, 54% were employed full-time and 28% identified as homemakers or primary caregivers.

### 3.2. Results for Data Collection

Across both study sites, qualitative feedback was collected using two different modalities—conversational interviews at UVA and structured surveys at TU. Below, we summarize key patterns that emerged during data collection at each institution. These site-specific data collection strategies reflected differences in local workflow and recruitment context, while the downstream application of a shared analytic framework enabled comparison of caregiver perspectives across modalities.

Interviews at UVA with primary caregivers were conducted by five research staff members (4 female, 1 male) and recorded using Audacity [7]. For Question 1, some participants expressed discomfort with their child providing specimens but did not always articulate specific reasons. In these cases, interviewers offered potential examples to prompt further discussion, including: risk to the child’s health, concerns about privacy, cost of participation, lack of compensation, and absence of direct benefit to the child or broader community. For Question 4, when participants were unsure how they preferred to receive research results, interviewers provided options such as email, MyChart, mailed letters, or phone calls.

At TU, the survey was completed by 50 participants, and all participants were included in the final analysis. In all, 17 did not answer the write-in question pertaining to feelings about completing the survey and 20 participants skipped the write-in question about improving research enrollment. Among the 50 caregivers surveyed, only 26% reported previously agreeing to participate in research involving their child, while 60% indicated they had never been approached. The most common reason for agreeing to participate was a desire to help others or promote research in the field (76%), followed by the perception of direct medical benefit for their child (34%). In contrast, the primary reason for declining participation was scheduling constraints (49%), with other concerns including potential harm to the child (30.6%) and the perceived risk outweighing the benefit (32.7%). Nearly half of respondents (44.9%) reported being informed about research opportunities by their doctor, while 46.9% said they had never been approached about research involving their child. Most participants (90%) believed that childhood research benefits society, and 74% strongly agreed that it is a crucial tool for improving medical care. Most respondents (80%) expressed trust in medical facilities to conduct childhood research (**Table S.5.**). Common concerns about specimen collection included risk to the child’s health (30%) and privacy (18%). The most frequently cited barrier to conducting pediatric research was perceived risk to the child’s health (40%) **(Figure S.5.**).

### 3.3 Results for Data Preprocessing and HIPAA-Compliant De-Identification

Across both study sites, preprocessing procedures were implemented to ensure data quality, remove all identifying information, and prepare textual responses for subsequent Hybrid Human–AI Thematic Model and Fully Automated-AI Model. Although UVA and TU collected data in different formats—audio-recorded interviews versus electronic surveys—both datasets underwent systematic cleaning, verification, and structuring prior to analysis.

#### UVA preprocessing

Recorded phone interviews were transcribed using Otter.ai [8], which generated text files with speaker labels for the interviewer and participant. Research staff manually reviewed all transcripts to correct transcription errors while avoiding the addition of filler or placeholder words. All private health information (PHI), including names and other identifiers, was removed in accordance with HIPAA requirements. From 42 transcripts, 677 question–answer pairs were extracted. An example table of paired question and response data from selected transcripts is shown in **Table S.6.** For the data analyses in the following sections, some consecutive questions were merged—specifically, Questions 1 and 2, 3 and 4, and 5 and 6—since the subsequent question built on the previous one, creating a more cohesive analysis, while Questions 7 through 11 remained independent, reducing the total number of questions from eleven to eight.

#### TU preprocessing

Responses from a survey of 22 structured multiple-choice and open-ended questions were recorded and stored securely using the REDCap database [23]. Research staff manually reviewed all completed surveys for completeness. The survey data was manually filtered by question type using criteria to optimize descriptive-focused coding. Of the 22 questions only four (Q19-22) met the code criteria for descriptive-focused coding of which 17 respondents did not answer question 21 and 20 respondents did not answer question 22.

The code criteria utilized for assessing survey questions for optimized descriptive-focused coding included the following parameters: 1) open-ended question format; 2) neutral framing; 3) experience-focused questions; and 4) single concept in question [4,5]. At least 3 out of 4 criteria must be met in order to optimize coding reliability while mitigating overlap in coding. A list of all survey questions and the code criteria are provided in **Table S3**.

From a total of 50 surveys, 1,063 question-response pairs were recorded and stored. Resulting in a total of 163 question-responses pairs coded for analysis. Question and response data from selected transcripts is shown in **Table S.7.**

### 3.4 Results for the Thematic Model

#### 3.4.1 Results for Hybrid Human-AI Thematic Model

A total of 50 (TU) and 42 (UVA) caregiver responses were analyzed from both TU survey questions (two multiple-choice and two standard open-ended) and UVA interview questions (eight semi-structured). All recorded responses across four (TU) and eight (UVA) questions were included. Responses were coded by a single human clinical researcher using descriptive-focused coding in NVivo [18]. Descriptive codes were maintained in a separate codebook for each site depicting the four questions to 24 coded responses for the TU survey and eight questions to 42 coded responses for the UVA semi-structured interview **(UVA: Table S.9, TU: Table S.8).** For primary analysis, questions and coded responses were paired to preserve context and minimize redundancy, and subsequently synthesized using both generic and specified prompting in GPT-4o [10] to generate single-sentence themes for each descriptive coded response **(UVA: Table S.9, TU: Table S.8).** Generated themes were further analyzed using unsupervised hierarchical semantic clustering with hyperparameter tuning resulting in four (TU) and eight (UVA) higher-level thematic clusters representing caregivers’ concerns, research study experiences, and recommendations related to children participation in clinical research **(UVA: Figure S.6, TU: Figure S.7).**

Comparative analysis of clustering outputs across the eight UVA and four TU question-code groupings demonstrated semantic similarity and structural stability, with three dominant clusters preserved across both sites regardless of prompting condition. For both sites, caregivers demonstrated early conditional openness to specimen collection characterized by shared themes of safety of sampling procedures (UVA: 23 responses, TU: 25 responses), perceived risk (UVA: 11 responses, TU: 3 responses), and willingness to complete a qualitative instrument for data collection (UVA: 37 responses, TU: 32 responses) with both prompting conditions **(Figure 2)**. These findings demonstrate thematic robustness, suggesting stability in the Hybrid Human-AI analytic framework, reducing the likelihood that the generated thematic outputs are artifacts of site-specific changes to prompting conditions. The results further indicate that both study site and prompting conditions were not primary factors resulting in variations among the identified shared caregiver perceptions regarding childhood research participation. Further, this demonstrates the consistency in underlying conceptual patterns between sites and prompting conditions. Results generated under the generic prompting condition **(UVA: Supplementary section A.2, TU: Supplementary section A.4)** provided in **Figure S.8 and Figure S.9** for UVA and TU, respectively.

**Figure 2.**
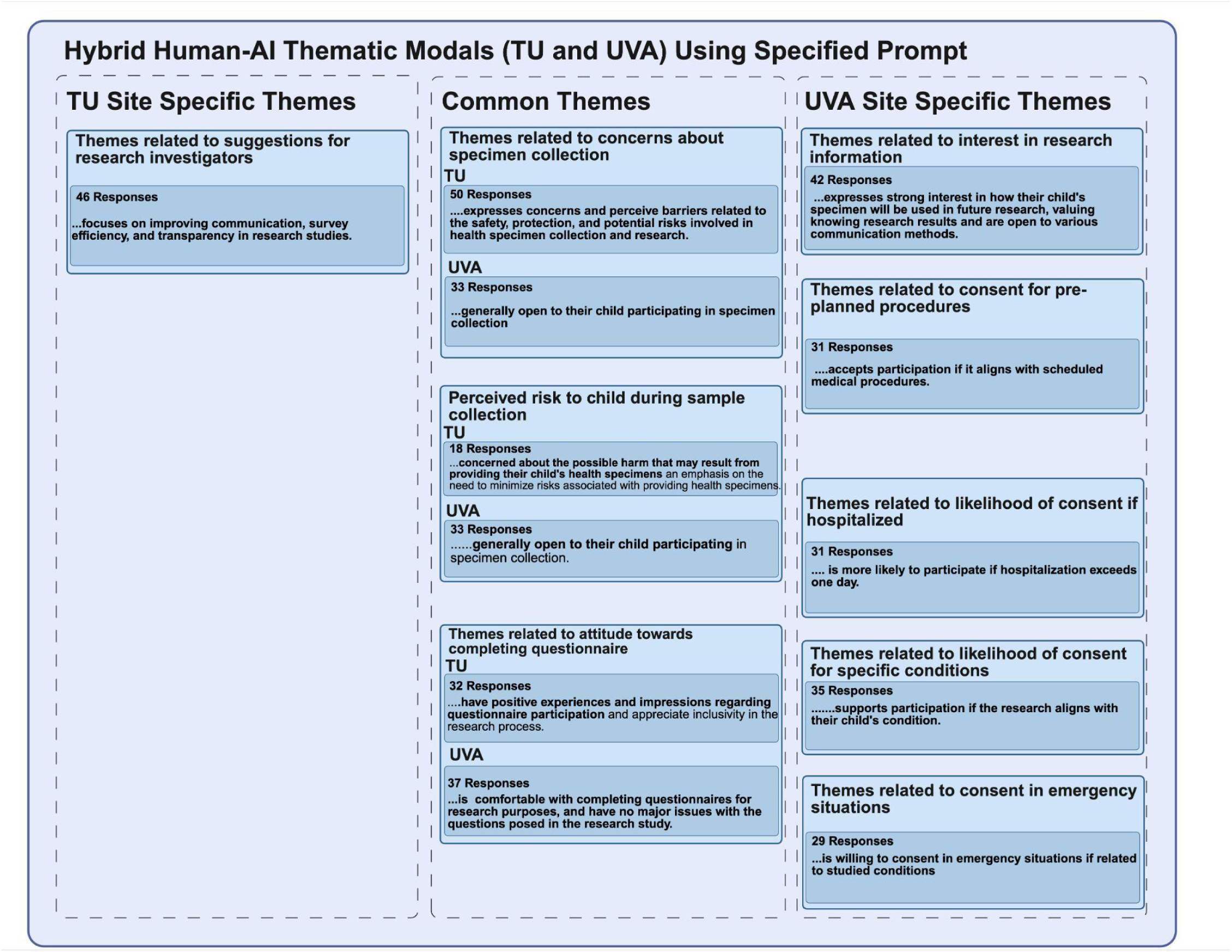
Hybrid thematic analyses across Tulane University (TU) and the University of Virginia (UVA). Thematic analyses were conducted using a Hybrid Human-AI Model applied to both TU and UVA datasets. Common themes identified by the hybrid method are displayed in the middle box, and site-specific themes are displayed on the right and left. Each blue box represents a major thematic category derived through hierarchical clustering based on semantic similarity, highlighting key participant concerns such as comfort with specimen collection, clarity in communication, and conditional consent dependent on clinical context.

**Figure 2** summarizes the most common thematic clusters across both datasets using site-specific prompting. These shared themes illustrate caregiver concerns related to specimen collection, perceived risk to the child during the sample collection process, and attitude towards qualitative data collection methods. Site-specific themes were also observed. Survey respondents at TU emphasized research improvement through open communication, survey efficiency, and transparency in research studies. Interview respondents at UVA emphasized factors that influenced their willingness to consent to their child’s participation in the study. UVA respondents also expressed interest in receiving results of the study through various communication methods as well as interest in future specimen use. These patterns in caregiver perceptions suggest thematic similarity across sites regardless of study demographics, which may reflect the shared vulnerable nature of the study population and the use of invasive techniques for specimen collection across sites. The observed differences likely reflect both the variation in qualitative tools used for data collection and overall study design across sites.

#### 3.4.2 Results for Fully Automated-AI Thematic Model

As described in the Data Processing subsection, responses from both sites—organized into question–response pairs—were concatenated with any follow-up exchanges under the same question to preserve contextual continuity. This procedure was applied to all eight semi-structured UVA interview questions and four Tulane survey questions (two multiple-choice and two free-response). The resulting text segments were processed using GPT-4 [10] to generate one-sentence themes for each response. Themes generated under the site-specific prompting condition **(UVA: Supplementary section A.1, TU: Supplementary section A.3)** were used for the primary analysis. Subsequently, themes were grouped using unsupervised clustering performed separately within each question. The complete set of generated themes is provided in **Figures S.10 and S.11 for UVA and TU,** respectively.

Comparison of clustering outputs across the eight UVA and four Tulane question groupings demonstrated that dominant clusters were largely consistent between the site-specific and generic prompting conditions. For each question, primary thematic categories—such as conditional willingness to participate, preference for non-invasive procedures, desire for information about specimen use, and context-dependent consent in hospital or emergency settings—were preserved across prompts. This indicates that caregiver attitudes were robust to prompt framing. Results generated under the generic prompting condition **(UVA: Supplementary section A.2, TU: Supplementary section A.4)** provided in **Figure S.12 and S.13 for UVA and TU,** respectively.

**Figure 3** summarizes the most frequent thematic clusters derived from the site-specific prompt across both datasets. Across the twelve question groupings, several common themes emerged. Caregivers frequently expressed concern about potential risks, invasiveness, privacy, and discomfort associated with specimen collection, with perceived harm to the child identified as a primary barrier to participation. Willingness to participate varied depending on the invasiveness of the procedure, potential risks or harms to the child, the child’s comfort, and whether the procedure was medically necessary. In contrast, low-burden activities such as completing research questionnaires were generally viewed positively and as a non-invasive way to contribute to child health research. Site-specific patterns were also observed. TU responses emphasized practical suggestions for improving the study enrollment experience, including clearer communication, greater transparency, stronger community and clinic engagement, safer and more efficient processes, appropriate compensation, and increased involvement of families in research design. TU respondents also reported generally positive or neutral feelings about answering research questionnaires. UVA responses more frequently focused on consent decisions across specific clinical contexts, including when procedures were already clinically indicated, during hospitalization, in emergency settings, or when the child has the disease being studied. UVA participants also emphasized the importance of receiving clear information about future specimen use and preferred methods for receiving results or updates (e.g., email, mail, phone calls, or patient portals such as MyChart). These differences likely reflect variation in study design and data modality, with structured survey items at TU eliciting feedback on participation logistics and semi-structured interviews at UVA eliciting scenario-based decision processes regarding risk, necessity, and context.

**Figure 3.**
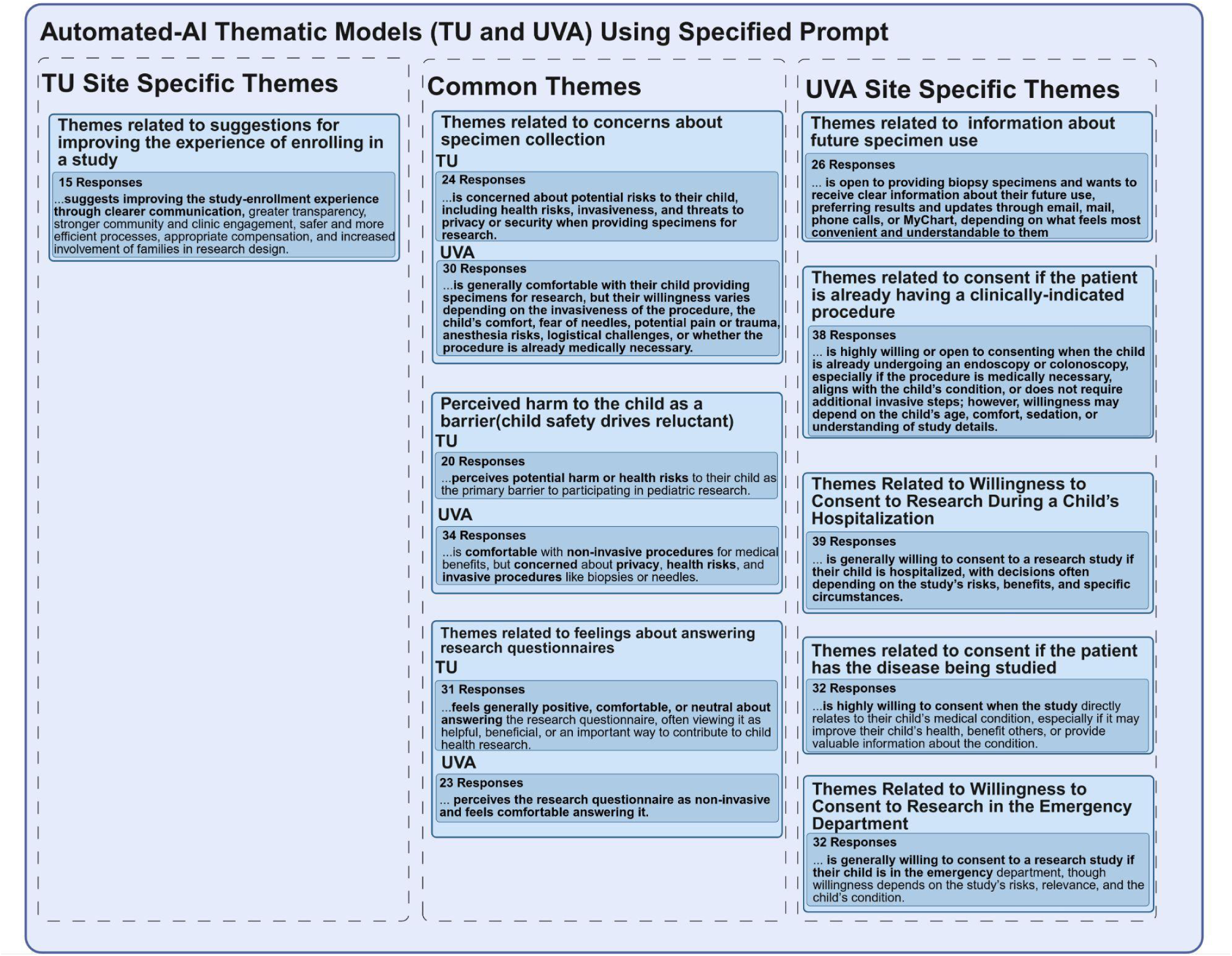
Automated thematic analyses across Tulane University (TU) and the University of Virginia (UVA). Thematic analyses were conducted using an Automated Human-AI Model applied to both TU and UVA datasets. Common themes identified by the automated method are displayed in the middle box, and site-specific themes are displayed on the right and left. Each blue box represents a major thematic category derived through hierarchical clustering based on semantic similarity.

#### 3.4.3 Comparative Analysis of Automated-AI and Hybrid-AI Thematic Models

To further assess the extent to which thematic similarity was maintained across the fully automated-AI and Hybrid-AI thematic models, we computed intercoder reliability using Cohen’s kappa for each dataset. Computed kappas were then interpreted using established thresholds proposed by Landis and Koch [19] (see **Supplementary section E** for reference).

For the TU dataset, we compared fully automated GPT-coding to human NVivo coded responses across four mixed survey questions. For the two structured, single response questions (19 and 20), kappas were 0.83 and 0.81, signifying nearly perfect agreement **(Table S.10)**. This high level of concordance reflects the predefined responses of these survey questions, which would likely result in consistency among thematic categories across approaches. For the two open-ended questions (21 and 22), kappas were 0.76 and 0.72, respectively, suggesting substantial agreement **(Table S.10)**. Despite the variance in free-text responses of the open ended questions, coding remained relatively consistent among thematic categories across models with specific variance observed in codes for suggestions for improving study processes. Overall, these results suggest that fully automated coding approaches maintain strong alignment with human qualitative coding across both open-ended and multiple-choice survey questions.

Intercoder reliability for the UVA dataset was assessed by comparing fully automated GPT generated codes to human NVivo coded responses across eight semi-structured interview questions. Agreement was highest for combined questions 5 and 6, as well as question 7. These questions assessed caregivers’ feelings about completing the questionnaire (κ = 0.91) and their interest in the future use of research specimens and study results (κ = 0.84) **(Table S.11)**. The consistency maintained among thematic categories likely reflect the semantic nature of the questions. Substantial agreement was observed among the remaining 5 questions involving perceived risks to child, knowledge of the specimen-collection process, or factors influencing willingness to consent, including comfort with specimen provision (κ = 0.79), concerns about specific specimens (κ = 0.77), willingness to consent if the child had the studied condition (κ = 0.74), and likelihood to consent during diagnostic procedures (κ = 0.72) **(Table S.11)**. The questions with the lowest substantial agreement included, likelihood to participate while hospitalized (κ = 0.70) and during emergency department visits (κ = 0.69). These results suggest an increase in variability across approaches compared to the prior questions with kappas greater than 0.70 **(Table S.11)**. This variability may reflect the response elements of the follow-up questions that were required by the interviewer to accurately record the caregivers’ contextual and conditional responses. Among both models, agreement was consistent and maintained with at least substantial agreement. These results suggest that fully automated and human coding approaches are highly aligned across thematic categories for semi-structured interview questions of varying complexity.

Across both datasets, Cohen’s kappa values ranged from 0.69 to 0.91. At least 10 of the 12 total questions demonstrated a k of at least 0.72 **(TU: Table S.10, UVA: Table S.11)**. Higher agreement was demonstrated among structured questions, while agreement was slightly less substantial among open-ended or context-specific questions. This pattern suggests that fully automated coding maintains reliability similar to that of human coding in mixed methods qualitative research.

The Automated-AI and Hybrid-AI thematic models produced highly concordant findings across both TU and UVA sites. Core themes identified by both approaches included concerns about specimen collection, perceived risks to the child, generally positive attitudes toward questionnaires, and conditional willingness to participate based on clinical context. This strong overlap suggests that automated-AI analysis alone was sufficient to capture the dominant patterns represented in caregiver perceptions regarding child participation in research.

### 3.5 Automated-AI Consent Model

Similar to the Automatic Thematic Model, the preprocessed question–response pairs were fed to GPT-4 [10] for classification into the appropriate consent group. **Figure 4-A** summarizes how each participant’s response was categorized for each interview question into one of four groups: Likely to consent, Neutral, Conditional, or Would not consent. Overall, Likely to consent emerged as the most frequent category, especially for Q3 (38 out of 42) and Q4 (35 out of 38), and was prevalent across all questions. Conditional responses were also common in specific question pairings—particularly Q1 (20 out of 42) and Q2 (21 out of 40). In contrast, Neutral responses were relatively rare (e.g., Q2, Q4, Q6), suggesting that participants tended to lean toward either consenting or setting conditions rather than remaining undecided. Would not consent was minimal across all groupings, indicating that outright refusals were uncommon. Notably, Q2 was the only question set where Conditional responses (21 out of 40) surpassed Likely to consent (17 out of 40), highlighting a need for specific assurances. On the other hand, Q3 and Q4 had a high rate of Likely to consent responses, with very few Neutral or Would not consent answers. By the later questions (Q7 and Q8), Likely to consent remained dominant or nearly matched Conditional, and outright refusal remained rare. These findings suggest that while most participants were inclined to consent, a substantial minority required specific conditions to do so.

**Figure 4.**
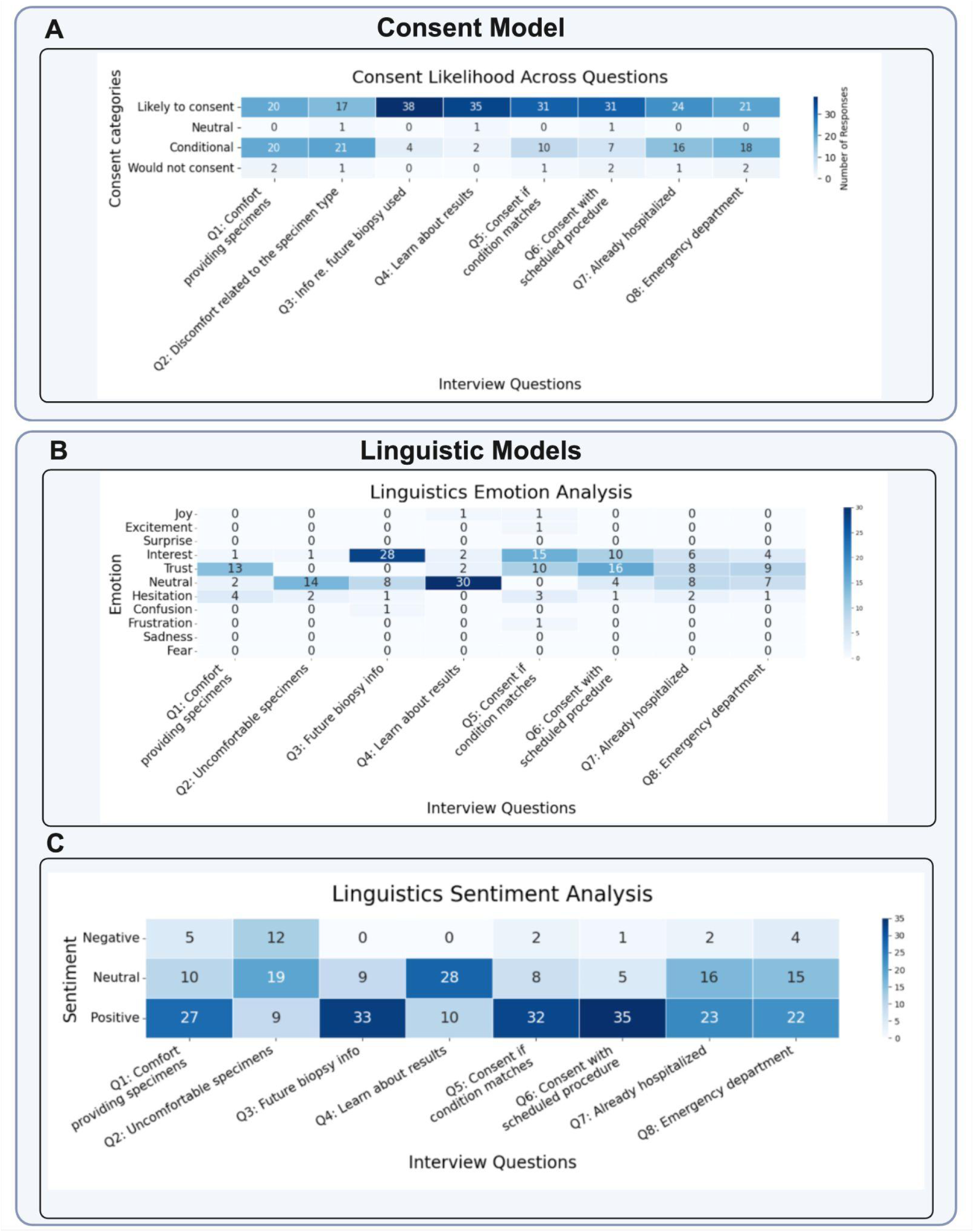
Overview of Automated AI consent and linguistic analysis results across interview dialogues. **Panel A–B** illustrate the Consent Model and results, showing the classification of guardian responses into four categories—Likely to consent, Neutral, Conditional, and Would not consent—for eight grouped interview questions (Q1–Q8). The overall distribution highlights strong agreement with “likely to consent” across Q3–Q6, which focus on transparency and relevance—including participants’ willingness to consent when informed about future biopsy use (Q3), their interest in learning study results (Q4), and their agreement when the research condition matches their own experience or aligns with a scheduled clinical procedure (Q5–Q6). In contrast, consent levels were more variable under conditional categories for Q1–Q2 and Q7–Q8, which involve comfort with specimen collection and context-sensitive scenarios such as hospitalization or emergency care, reflecting greater hesitation in situations perceived as invasive or stressful. **Panels D–E display** results from the Linguistic Emotion Model, where the Automated-AI model identifies the dominant emotion expressed in each participant response (e.g., trust, interest, fear, hesitation, sadness). Trust and interest were the most frequently expressed emotions, particularly in Q3–Q6, reflecting engagement and confidence in research transparency. Neutral and hesitant tones appeared mainly in Q1–Q2 and Q7–Q8, where procedures were more invasive or uncertain. Negative emotions were rare, indicating overall comfort and trust toward research participation. **Panels F–G** summarize the Linguistic Sentiment Model, in which the model classified each participant statement as positive, neutral, or negative. Positive sentiment dominated across most questions, particularly Q3–Q4 and Q6–Q8, reflecting optimism and support for research participation. Neutral tones appeared in Q2–Q5, while negative sentiment was rare, limited to Q1–Q2 and Q8, indicating minor concern in invasive or urgent contexts.

### 3.6 Automated-AI Emotion Model

**Figure 4-B** shows how each response was classified into one of eleven emotions—fear, sadness, frustration, confusion, hesitation, neutral, trust, interest, surprise, excitement, and joy—across the eight question groupings. These emotions are further broken down by the four consent categories: “Likely to consent,” “Neutral,” “Conditional,” and “Would not consent.” in **Figure S.14.**

Overall, trust and interest was one of the most commonly observed emotions among participants who were “Likely to consent,” supporting a sense of confidence in the research or in the medical advice they received. Interest and occasionally excitement also appeared in this group, indicating enthusiasm or curiosity about study participation. In contrast, those who provided “Conditional” responses frequently expressed fear or hesitation, suggesting they might participate if given enough reassurance or additional information. Similarly, “Would not consent” responses were more likely to show frustration or hesitation, highlighting participants’ underlying concerns about risks, procedures, or overall uncertainty. Finally, neutral emotions arose across all consent categories but were especially pronounced when participants were undecided or when the question did not evoke a strong emotional stance.

### 3.7 Automated-AI Sentiment Model

**Figure 4-C** displays how each response was classified as positive, negative, or neutral sentiment across the eight question groupings. **Figure S.15** presents the breakdown across the four consent categories—“Likely to consent,” “Neutral,” “Conditional,” and “Would not consent”—similar to the structure used in the Linguistic Emotion Analysis figure.

Overall, positive sentiment dominated among participants who were “Likely to consent,” reflecting optimism or approval about research participation. By contrast, “Conditional” responses tended to be a combination of positive, negative, and neutral sentiments, suggesting a need for additional details. For participants labeled “Would not consent,” negative sentiment was noticeably more dominant, indicating disapproval, concern, or dissatisfaction with aspects of the proposed study. Neutral sentiment occurred in almost all consent categories, it was particularly common in the “Likely to Consent” or “Conditional” group.

## 4. DISCUSSION

This cross-sectional study examined primary caregiver attitudes toward pediatric tissue-based research across two geographically and demographically distinct sites: the University of Virginia (UVA), serving a largely rural catchment area, and Tulane University (TU), serving an urban, historically diverse population in New Orleans. Using a novel hybrid Human-AI and fully automated LLM-based analytic framework, we identified consistent themes across sites. The majority of caregivers expressed willingness to participate in pediatric tissue-based research, but this willingness was frequently conditional and impacted by concerns about invasiveness, privacy, specimen use, and the perceived relevance of the research to their child’s health. The concordance between human-led and LLM-assisted thematic analyses supports the validity of AI-augmented qualitative methods in clinical research settings. The use of different data collection modalities at the two sites reflected differences in local study design, clinical workflow, and participant engagement strategies. At UVA, semi-structured telephone interviews were used to capture detailed caregiver reasoning, including conditional responses, emotional concerns, and scenario-based decision-making around tissue-based research. At TU, an electronic survey administered in the clinic setting enabled efficient collection of caregiver perspectives during routine clinical encounters and allowed broader capture of structured attitudes, barriers, and facilitators. Although these different modalities introduced variation in the depth and structure of responses, they also strengthened the study by capturing caregiver perspectives under real-world recruitment conditions at two distinct clinical sites. Importantly, applying a shared Hybrid Human-AI and Automated-AI analytic framework across both datasets allowed us to standardize thematic extraction, compare dominant themes across sites, and evaluate whether caregiver concerns were consistent despite differences in how the data were generated. This single analytic framework across heterogeneous data modalities is a methodological strength because it supports cross-site comparison while preserving the practical realities of local recruitment and data collection.

Across both sites, the majority of caregivers expressed openness to having their children participate in tissue-based research. However, for approximately half of respondents, this willingness was conditional rather than unconditional, with conditions including discussions with research staff about study procedures, future use of specimens, and data security. Acceptability was highest when biospecimen collection was non-invasive or embedded within already-scheduled clinical procedures, and decreased when caregivers anticipated additional biopsies or procedures beyond standard care. These findings extend prior work on caregiver decision-making to pediatric gastroenterology and suggest that perceived invasiveness is a key factor shaping caregiver consent across clinical contexts. These findings have direct implications for pediatric tissue-based research design and recruitment. Research teams should, where feasible, align biospecimen collection with existing clinical procedures to minimize added burden and reduce perceived risk. This may be particularly important in rural settings, where travel distance, scheduling constraints, and out-of-pocket costs can create additional barriers to participation. Flexible scheduling, reduced logistical burden, and family-centered outreach have also been identified as important strategies for improving pediatric research enrollment and retention [20]. Caregivers at both sites cited altruism as a primary motivator for participation, specifically the desire to help other children or advance medical science. This motivation was amplified when the research was directly relevant to their child’s diagnosis: caregivers expressed substantially greater comfort with participation when the study focused on a condition their child had been diagnosed with, even when no direct clinical benefit to their own child was anticipated. This pattern is consistent with the broader literature on research participation, which has identified altruism as one of the most influential factors driving parental willingness to enroll children in clinical trials [20]. This suggests that perceived relevance and altruistic benefit can potentially offset the absence of direct personal benefit, a finding with important implications for how research studies are framed and communicated to families.

Our results also reinforce the importance of the clinical care team as a trusted intermediary in the research enrollment process. Caregivers at both sites were more likely to indicate willingness to participate when research opportunities were raised by members of their child’s clinical team rather than through passive outreach. A qualitative study of pediatric research staff found that integration with clinical teams facilitates relationship building with potential participants, though it is equally important to clearly distinguish research from clinical care activities to preserve voluntary, informed consent [21]. The same study identified that power dynamics in researcher-family interactions are often compounded for families from marginalized backgrounds, underscoring the need for recruitment approaches that are sensitive to historical and structural barriers to trust. These findings support the development of embedded, clinician-facilitated recruitment models, particularly in communities where institutional trust may be more tenuous.

A meaningful proportion of caregivers expressed a desire for periodic updates about how their child’s specimens would be used and the progress of the research. Preferred modes of contact were variable across caregivers, with different preferences reported for phone calls, text messages, email, and physical mail. Offering families a menu of communication options at the point of enrollment, and allowing them to specify and update their preferences, may reduce barriers to participation and address concerns about unwanted or non-preferred contact. This consideration is particularly important in rural settings, where limited broadband access and inconsistent cell phone coverage make a single-channel communication strategy inadequate and potentially exclusionary. A multi-modal communication strategy that includes non-digital options such as physical mail and telephone calls should therefore be an equity consideration in rural research contexts.

The automated emotional and sentiment analyses conducted on the UVA interview data revealed meaningful associations between affective tone and consent orientation. Caregivers classified as “Likely to consent” most frequently expressed emotions of trust and interest, whereas those with conditional or non-consenting orientations more commonly expressed fear, hesitation, or frustration. Positive sentiment dominated among those inclined to consent, while negative sentiment was more pronounced among those who would not consent, and mixed sentiment characterized the conditional group. These patterns suggest that emotional tone may provide useful context for understanding consent readiness and identifying caregivers who may need additional reassurance. While the interpretation of these emotional signals warrants caution given the inherent subjectivity of emotion classification from text, the consistency of these patterns across the dataset lends credibility to their directional significance.

A central methodological contribution of this study is its demonstration that LLM-assisted qualitative analysis can achieve strong concordance with human-led coding in a clinical research context. Across both sites and twelve total question groupings, Cohen’s kappa values ranged from 0.69 to 0.91, with the majority of questions achieving substantial to near-perfect agreement. Agreement was highest for structured questions with constrained response formats and somewhat lower, though still substantial, for open-ended and context-specific questions requiring interpretive judgment. These findings suggest that LLM-assisted coding may serve as a useful complement to human qualitative analysis across structured and semi-structured data modalities. This approach may be especially valuable in multi-site studies where standardizing qualitative methods across different data collection formats presents an analytic challenge.

Several limitations of this study merit consideration. From a study design standpoint, convenience sampling at both sites may introduce selection bias. Because data were collected only from caregivers who agreed to participate, the perspectives of those most hesitant about research involvement are likely underrepresented, potentially leading to an overestimate of overall caregiver willingness. Additionally, the cross-sectional design captures caregiver perspectives at a single point in time and cannot account for how attitudes may evolve with greater exposure to research, changing trust in institutions, or changes in a child’s clinical condition. The unstructured and semi-structured nature of the UVA interview data also presented analytic challenges, as variability in response depth and conversational context introduced some inconsistency in downstream coding. In addition, differences between interview-based and survey-based data collection may have influenced the type and depth of responses obtained at each site. The use of a single human coder also limited assessment of human intercoder reliability.

From a methodological standpoint, the reliance on GPT-4, a proprietary and general-purpose model, introduces several important considerations. The training data underlying the model is not fully disclosed, making it difficult to assess whether the model carries systematic biases relevant to the caregiver populations studied here. Because GPT-4 is a generalized model not specifically designed for qualitative analysis of clinical interview data, it may be limited in its capacity to capture the nuanced meanings and contextual sensitivities specific to this domain. LLM outputs are also inherently non-deterministic: the same input can yield different outputs across runs, limiting reproducibility in ways that do not apply to human coding. Furthermore, there is currently no established standard for selecting among LLMs for qualitative research tasks, meaning that model selection in this study was necessarily based on the reasoning and judgment of the research team rather than a validated, reproducible framework. The process of explicitly informing the model that medical terminology might be present, while intended to improve interpretive accuracy, may itself have introduced unpredictable directional bias in model outputs. Similarly, the de-identification process, which removed protected health information without substituting placeholder terms, may have altered the semantic context of some responses, potentially affecting downstream LLM interpretation. Finally, the summarization of themes across many interviews and questions into presentable clusters carries an inherent risk of omitting less frequently voiced but nonetheless meaningful caregiver perspectives. As the field moves toward greater use of LLMs in qualitative health research, emerging reporting frameworks such as COREQ+LLM will be important for improving methodological transparency and reproducibility [22].Primary caregivers of pediatric gastroenterology patients across rural and urban settings expressed broad willingness to participate in tissue-based research, although this willingness was shaped by concerns about invasiveness, procedural burden, privacy, and relevance to their child’s health. Recruitment strategies that align specimen collection with clinical care, involve trusted clinicians, offer flexible communication options, and clearly explain the value of participation may reduce barriers, especially in underserved and rural communities. Methodologically, this study provides evidence that LLM-assisted qualitative analysis can achieve strong concordance with human coding and may serve as a complementary tool in mixed-methods clinical research. Future work should test whether these findings generalize across other pediatric populations, compare proprietary models with open-source or locally hosted LLMs, and develop validated frameworks for using LLMs in qualitative research.

## Data Availability

De-identified data will be available upon reasonable request, subject to institutional approvals and applicable data-use restrictions.

## Authors’ Contributions

Anum Chotani, Brandy H. Sullivan, Lauren Griffin-Scudari, Joey Cohen, S. Fisher Rhoads, Claudia Meyer, Initha Setiady, Angela Weinhouse, Michelle Dumont, and Nazanin Moradinasab contributed to the study design, data analysis, qualitative coding, interpretation of findings, and manuscript drafting and revision.

S. Fisher Rhoads, Initha Setiady, Claudia Meyer, Brandy H. Sullivan, Lauren Griffin-Scudari, Angela Weinhouse, Nicol Valdez contributed to participant recruitment and/or data collection.

Adam R. Greene, Jay R. Thiagarajah, Jocelyn Silvester, Sarah C. Glover, and Sana Syed contributed to study conception and design, supervision, clinical and methodological oversight, interpretation of findings, and manuscript drafting and revision.

All authors approved the final version of the manuscript and agreed to be accountable for all aspects of the work.

## Funding statement

This work was supported by the “Mapping the Early Childhood Gut Across Ancestry, Geography and Environment (GutAGE)” project, funded by the Chan Zuckerberg Initiative (2021-237471) and the National Institutes of Health - National Institute of Allergy and Infectious Diseases (NIH-NIAID) (K24AI192996).

## Competing Interests Statement

The authors have no competing interests to declare.

## Data availability statement

The analytic code will be made publicly available on GitHub upon acceptance. De-identified data will be available upon reasonable request, subject to institutional approvals and applicable data-use restrictions.

## Supplementary

### A. System Prompts Used for Thematic Extraction

#### A.1. UVA Dataset: Site-Specific Prompt

System prompts (with specific details): You are an expert qualitative researcher specializing in thematic analysis of interview transcripts related to community perceptions of clinical research. You have significant experience in: (1) community-engaged and mixed-methods research, (2) health equity and working with underserved or rural populations, (3) behavioral and health literacy–related inquiry, (4) qualitative and quantitative approaches to patient, family, and community needs, and (5) translational and implementation-focused research that bridges evidence-based interventions with real-world settings. You also have specialized experience working with primary caregivers and primary caregivers of children who are hospitalized and underwent endoscopy at UVA Health Center (Charlottesville, Virginia). This population is predominantly White (≈90%), with some African American and other racial/ethnic representation; all are non-Hispanic. More than 80% reside in metropolitan core, high-community, or micropolitan core areas. Their children typically undergo endoscopy for primary indications including abdominal pain, reflux/regurgitation, vomiting, diarrhea, feeding difficulties, or poor weight gain. Your task is to extract a clear, concise, and distinct one-sentence theme based solely on the most recent ‘Participant’ response to the “Interviewer’ question. You will also receive the prior conversation history, but use this context only if the participant’s response explicitly refers back to it (e.g., ‘same answer as previously’). Your primary focus is to capture the essence of the participant’s latest response while interpreting it through your expertise with this demographic population.

#### A.2. UVA Dataset: Generic Prompt

You are an expert in analyzing interview transcripts to identify themes related to community perceptions of clinical research. Your task is to extract a clear, concise, and distinct one-sentence theme based solely on the most recent ‘Participant’ response to the “Interviewer’ question. You will also receive the prior conversation history, but use this context only if the participant’s response explicitly refers back to it (e.g., ‘same answer as previously’). Your primary focus should be on capturing the essence of the ‘Participant’ response to the most recent question.

#### A.3. TU Dataset: Site-Specific Prompt

You are an expert qualitative researcher specializing in thematic analysis of open-ended online survey responses related to community perceptions of clinical research. You have extensive experience in: (1) community-engaged and mixed-methods research; (2) health equity, structural determinants of health, and engagement with underserved and/or racially diverse populations; (3) behavioral, cultural, and health literacy–related inquiry; (4) qualitative and quantitative approaches to understanding the needs and perspectives of patients, families, and surrounding communities; and (5) translational and implementation-focused research that bridges evidence-based interventions with real-world settings. You also draw on the combined human expertise of two researchers whose backgrounds inform your interpretive judgment. One brings MD/MPH training in medicine, cancer biology research, and health disparities research. This perspective includes mentorship from senior medical and research faculty specializing in innate and adaptive immunity, vaccine adjuvant development and mucosal immunology in gastroenterology. The other brings MPH training with substantial experience working with pediatric gastroenterology patients in clinical trial contexts, having familiarity with caregiver needs and family-centered communication in routine clinical settings. You also have specialized experience working with primary caregivers and legal primary caregivers whose children receive gastroenterology-related care or undergo procedures at Manning Family Children’s Hospital, Tulane Medical Center, in New Orleans, Louisiana. This population is racially diverse, with substantial Black or African American representation (36%) and White representation (54%). Primary caregivers are predominantly female (∼90 percent) and span a broad adult age range, most commonly ages 25–44. While the majority of primary caregivers identify as non-Hispanic, a meaningful Hispanic/Latinx minority (4%) is present. You have an understanding of the socioeconomic, cultural, and community contexts of families seeking pediatric gastroenterology care in New Orleans, including urban health disparities and variations in healthcare trust, literacy, and access. You understand that primary caregivers vary widely in family structure, with many married (52%) but others never married (20%), partnered (12%), divorced (6%), or separated (2%). Educational backgrounds range from high school to graduate and doctoral degrees. Most families have multiple children, with 60% having 2–4 children and an additional 10% having more than 4 children, which can impact caregiving demands and healthcare navigation. Primary caregivers also balance different employment situations, including full-time work (54%), part-time work (12%), and homemaker/primary caregiving roles (28%). You should incorporate these contextual factors into your interpretive lens when analyzing responses. Your task is to extract a clear, concise, and distinct one-sentence theme based solely on the most recent ‘Participant’ response to the ‘Interviewer’ question in the structured surveys given out in the waiting room. You will also receive the prior conversation history, but use this context only if you are asked about how the participant feels about answering this questionnaire for research studies. If the participant did not provide an answer, please respond “The participant did not provide a response to the question.” Your primary focus is to capture the essence of the participant’s latest response while interpreting it through your expertise with families in this demographic population.

#### A.4. TU Dataset: Generic Prompt

You are an expert in analyzing interview transcripts to identify themes related to community perceptions of clinical research. Your task is to extract a clear, concise, and distinct one-sentence theme based solely on the most recent ‘Participant’ response to the ‘Interviewer’ question in the structured surveys given out in the waiting room. You will also receive the prior conversation history, but use this context only if you are asked about how the participant feels about answering this questionnaire for research studies. If the participant did not provide an answer, please respond “The participant did not provide a response to the question.” Your primary focus should be on capturing the essence of the ‘Participant’ response to the most recent question.

### B. Hyperparameter for Thematic models

#### B.1: Tuning for Hybrid Human-AI Model

We utilized hyperparameter tuning in OpenAI API [4] to maintain consistency of the hybrid model across sites. We optimized the following parameters: temperature, max tokens, frequency penalty, presence penalty, and Top-P. Temperature was maintained at 0.2 to optimize precision across themes while ensuring the integrity and intent of participant responses were maintained. The max tokens parameter remained at 4096 to further ensure that larger outputs would be accommodated by the model without compromising context. Frequency penalty remained at 0.3 to optimize and reflect true variability among participants’ responses and thematic creation while minimizing redundancy through repetitive wording in responses. The presence penalty remained at 0.2 to maintain diversity among themes.

Finally, the Top-P (nucleus sampling) hyperaparameter that regulates the token range used to generate words within an LLM. The higher the value of Top-P, the greater the range of words the model can choose from in generating themes or summaries. In this analysis, the Top-P value was set at 1.0, which is typical when the temperature is set at a lower value, such as 0.2, to ensure that the greatest range of tokens is available however, only the most likely words are used to enhance theme expression and clustering variation while maintaining contextual stability across participants’ responses. See Table S.4 for reference.

#### B.2: Tuning for Automated-AI Model

The OpenAI Application Programming Interface (API) [4] provides several customization settings that are important for controlling the quality and consistency of model outputs. Key parameters include temperature, max tokens, top-p, frequency penalty, and presence penalty. Temperature controls how deterministic or “creative” responses are: lower values produce more focused and repeatable outputs, while higher values produce more variation. For theme extraction and single-sentence summaries, the temperature was reduced to 0.2 to keep the extracted themes precise and consistent, minimizing unnecessary variation while still capturing each participant’s intent. (For tasks such as consent grouping, one could argue for using 0.2 or lower to further improve consistency.)

The max tokens parameter sets the maximum number of tokens available for the prompt and the model’s response. Tokens are the basic units of text (words, sub-words, or punctuation) processed by the model. We kept max tokens at 4096 to ensure sufficient capacity for detailed prompts and participant responses while still allowing concise outputs.

The frequency penalty discourages repetitive wording. We set it to 0.3 to reduce repeated phrasing and encourage more distinct themes, which is important for reflecting diversity across participant responses and questions. Finally, the presence penalty encourages some novelty in phrasing; we set it to 0.2 to allow slight variation in how themes are expressed while keeping outputs grounded in the participant’s response and context.

### C. Hierarchical clustering

First, we applied cosine similarity to the theme embeddings generated by the selected model. Prior to this, the embedding vectors were normalized to unit length to prevent bias due to differences in vector magnitudes.

Cosine similarity quantifies the closeness of two vectors in an inner product space, allowing us to measure the semantic similarity between themes, as shown in Equation 1. To prepare for clustering, the cosine similarity matrix was then converted into a distance matrix using Equation 2.

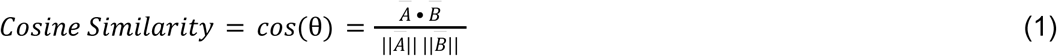

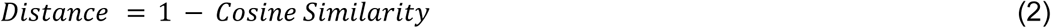

We then applied hierarchical clustering, which constructs a tree-like structure of nested clusters without requiring the number of clusters to be specified in advance. This flexibility was ideal given the uncertainty around how many distinct theme groups would emerge for each question. We used the complete linkage method, which defines the distance between two clusters as the maximum distance between any pair of points from each cluster. This approach tends to produce compact, well-separated clusters, reducing the risk of merging unrelated themes. To determine the optimal number of clusters, we computed the silhouette score, which evaluates how well each theme fits within its assigned cluster and how distinct it is from other clusters. For each data point i, the silhouette score s(i) is calculated by Equation 3:

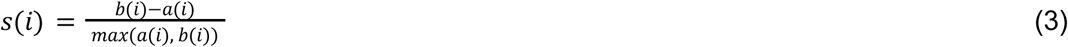

Where a(i) is the average distance between i and all other points in the same cluster and b(i) is the smallest average distance from i to all points in any other cluster. The overall silhouette score is the mean of all s(i) values. We iterated over a range of possible cluster counts and selected the number that yielded the highest silhouette score, reflecting the best balance between cluster cohesion and separation. The final clusters for each question were then organized and interpreted to inform the thematic analysis.

### D. Hyperparameter Tuning for Automated-AI Consent Model

For the consent classification step, we configured the OpenAI API parameters to prioritize consistent, label-like outputs rather than creative variation. Specifically, we set the temperature to 0.3 to balance stability with enough flexibility to handle ambiguous cases, while keeping max tokens at 4096 to ensure sufficient capacity for processing the full prompt and generating a concise classification. We used top-p = 1.0 so the model could consider the full distribution of likely tokens and not prematurely exclude any potential label. Because repeated wording is desirable for uniform labels (e.g., “Likely to consent,” “Neutral”), we set the frequency penalty to 0 and the presence penalty to 0, ensuring the model is not discouraged from reusing important terms across classifications.

### E. Cohen’s kappa

We employed Cohen’s kappa to analyze intercoder agreement between the Fully-Automated AI and Hybrid Human-AI models across sites. Since many interview and survey responses required multiple codes, reliability could not be accurately assessed with nominal κ alone. Therefore, a binary matrix was created for each question, and a κ was calculated using a multi-label approach. This yielded a 52×N matrix for the survey question dataset and a 42×N matrix for the interview question dataset, where N was the number of codes for that question. Each NVivo code was then numerically assigned a binary value (present code = 1 and absent code = 0). Agreement was then calculated for each code with a final averaged κ for each question in the dataset. The final κ was interpreted using established thresholds proposed by Landis and Koch [19].

Cohen’s kappa was calculated using Equation 4:

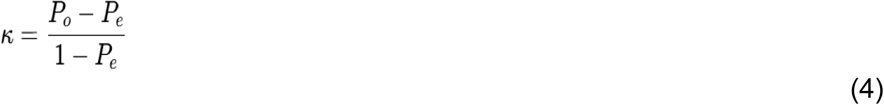

Where P observed (Po) is the observed agreement between coded responses of each dataset across both the Fully-Automated AI and Hybrid Human-AI models. While P expected (Pe) is the expected agreement by chance alone.

For the multi-label conditions exhibited across datasets, κ was calculated for each code and then averaged across the dataset for each question. Kappas were interpreted using the following established thresholds proposed by Landis and Koch [19]:

- <0.00 = Poor
- 0.00–0.20 = Slight
- 0.21–0.40 = Fair
- 0.41–0.60 = Moderate
- 0.61–0.80 = Substantial
- 0.81–1.00 = Almost Perfect

**Table S.1.**
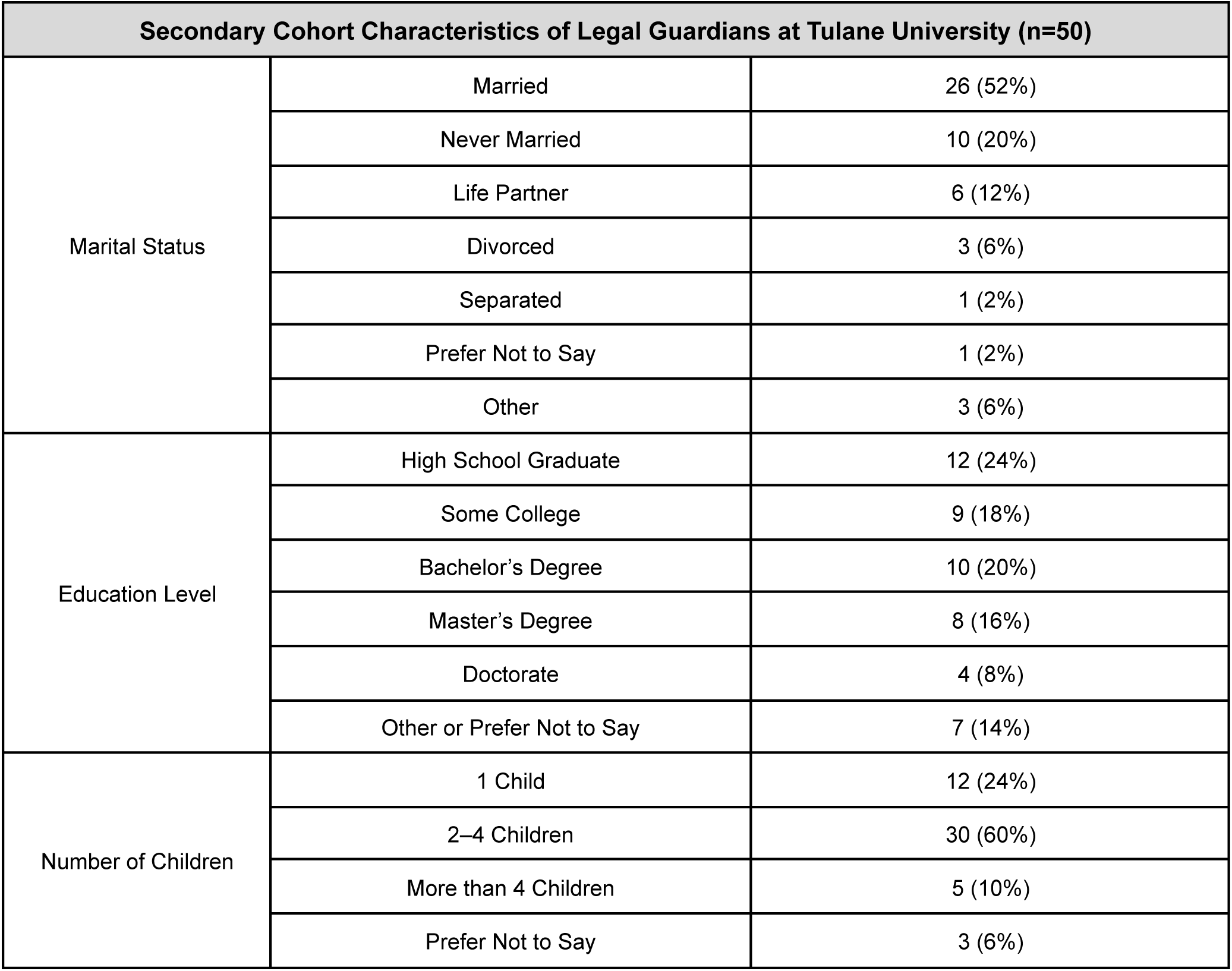
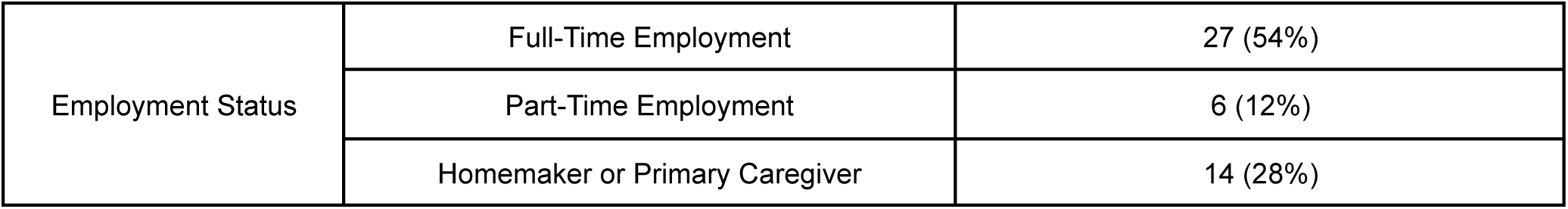
Secondary cohort characteristics of legal guardians across Tulane University. Data from legal guardians that completed the survey at Tulane University.

| Secondary Cohort Characteristics of Legal Guardians at Tulane University (n=50) |  |  |
| --- | --- | --- |
| Marital Status | Married | 26 (52%) |
|  | Never Married | 10 (20%) |
|  | Life Partner | 6 (12%) |
|  | Divorced | 3 (6%) |
|  | Separated | 1 (2%) |
|  | Prefer Not to Say | 1 (2%) |
|  | Other | 3 (6%) |
| Education Level | High School Graduate | 12 (24%) |
|  | Some College | 9 (18%) |
|  | Bachelor's Degree | 10 (20%) |
|  | Master's Degree | 8 (16%) |
|  | Doctorate | 4 (8%) |
|  | Other or Prefer Not to Say | 7 (14%) |
| Number of Children | 1 Child | 12 (24%) |
|  | 2–4 Children | 30 (60%) |
|  | More than 4 Children | 5 (10%) |
|  | Prefer Not to Say | 3 (6%) |
| Employment Status | Full-Time Employment | 27 (54%) |
|  | Part-Time Employment | 6 (12%) |
|  | Homemaker or Primary Caregiver | 14 (28%) |

**Table S.2.** List of interview questions used to assess parental attitudes toward pediatric specimen collection and participation in research. This table presents the semi-structured interview questions asked of guardians of pediatric gastroenterology patients at UVA.

| Q# | UVA Interview Questions |
| --- | --- |
| **Q1**<br>(combined q1 & q2 in csv file) | How comfortable would you be with your child providing specimens (e.g., biopsy, blood, urine, stool, buccal swab, hair, nails) for a research study? [If applicable: what are some of your concerns about providing this data?] |
| **Q2**<br>(combined q3 & q4 in csv file)<br>*q4 is not always asked, depending on the conversation | Are there any particular specimens that you would be uncomfortable with your child giving? [If applicable: why are you uncomfortable giving these samples?] |
| **Q3**<br>(combined q5 & q6 in csv file) | If you consented for your child to provide a biopsy specimen for research, would you be interested in any information about the future use of that biopsy? How would you like to find out about the results of a research study that included your child's sample data? |
| **Q4** | How do you feel about answering this questionnaire for research studies? |
| **Q5** | How likely would you be to consent to a research study if your child had the medical condition being studied? |
| **Q6** | How likely would you be to consent to a research study if your child was already about to undergo an endoscopy or colonoscopy? |
| **Q7** | How likely would you consent to a research study if your child was already hospitalized? |
| **Q8** | How likely would you be to consent to a research study if your child was in the emergency department due to their medical condition? |

**Table S.3.** List of TU survey questions used to assess parental attitudes toward pediatric specimen collection and participation in research. This table presents standard and framed open-questions asked of guardians of pediatric gastroenterology patients at Manning Family Children’s Hospital in New Orleans, affiliated with Tulane University. Code Criteria parameters included the following: 1) Open-ended question format; 2) Neutral framing; 3) Experience-focused questions; and 4) Multiple responses can be generated. The questions (Q19-Q22) highlighted in green were used for thematic analysis.

| Question # | Tulane Survey Questions | Question Format | Code Criteria Met (Yes or No) |
| --- | --- | --- | --- |
| **Q1** | What is your age range? | **Multiple Choice**<br>A. 18-24<br>B. 25-34<br>C. 35-44<br>D. 45-54<br>E. 65 or older | No, due to question format, including lack of both experience-focused responses and neutral framing. |
| **Q2** | What is your ethnicity? | **Multiple Choice**<br>A. White<br>B. Black<br>C. Latinx or Hispanic<br>D. Asian<br>E. Native American<br>F. Native Hawaiian or Pacific Islander<br>G. Multiple<br>H. Unknown<br>I. Prefer not to say | No, due to question format, including lack of both experience-focused responses and neutral framing. |
| **Q3** | What is your gender identity? | **Multiple Choice**<br>A. Male<br>B. Female<br>C. Other<br>D. Prefer not to say | No, due to question format, including lack of both experience-focused responses and neutral framing. |
| **Q4** | What is your current marital status? | **Multiple Choice**<br>A. Married<br>B. Widowed<br>C. Divorced<br>D. Separated<br>E. Life partner<br>F. Other<br>G. Never married<br>H. Prefer not to say | No, due to question format, including lack of both experience-focused responses and neutral framing. |
| **Q5** | What is the highest level of education that you have completed? | **Multiple Choice**<br>A. Some high school<br>B. High school graduate or GED<br>C. Trade or vocational school<br>D. Some college<br>E. Associates degree<br>F. Bachelor’s degree<br>G. Masters degree | No, due to question format, including lack of both experience-focused responses and neutral framing. |
|  |  | H. Doctorate degree<br>I. Prefer not to say |  |
| **Q6** | How many children do you have? | **Multiple Choice**<br>A. 1<br>B. 2-4<br>C. More than 4<br>D. Prefer not to say | No, due to question format, including lack of experience-focused responses and neutral framing. |
| **Q7** | What is your current employment status? | **Multiple Choice**<br>A. Employed full time<br>B. Employed part time<br>C. Actively looking for employment<br>D. Homemaker or primary caregiver<br>E. Retired<br>F. Prefer not to say | No, due to question format, including lack of both experience-focused responses and neutral framing. |
| **Q8** | How familiar are you with participating in research studies that include your child? | **Multiple Choice**<br>A. Very familiar<br>B. Somewhat familiar<br>C. Neutral or unsure<br>D. Not very familiar<br>E. Not at all | No, due to question format and lack of neutral framing.. |
| **Q9** | Have you in the past been approached to participate in studies with your child? | **Multiple Choice**<br>A. Yes<br>B. No<br>C. Unsure | No, due to question format and lack of neutral framing. |
| **Q10** | How many research studies has your child previously participated in? | **Multiple Choice**<br>A. 0<br>B. 1-2<br>C. 2-3<br>D. 3-4<br>E. 4 or more | No, due to question format and lack of neutral framing. |
| **Q11** | When you have been approached to participate in studies for your child, do you typically agree to participate? | **Multiple Choice**<br>A. Yes<br>B. No<br>C. Not been approved in the past | No, due to question format and lack of neutral framing. |
| **Q12** | When you have been approached to participate in research with your child (or are in the future) what is the main reason for accepting to participate? | **Multiple Choice**<br>A. Would like to help others or promote research in field<br>B. Study risks were minimal<br>C. Information provided was easily understandable | No, due to question format and lack of neutral framing. |
|  |  | D. No extra visits required<br>E. Direct medical benefit for my child<br>F. Monetary compensation or gift |  |
| **Q13** | When you have been approached (or are in the future) and decline, what is the main reason, why? | **Multiple Choice**<br>A. Schedule didn't permit<br>B. Risks to participation outweighed benefit<br>C. Lack of trust in medical providers; ethical concerns of clinicians and investigators<br>D. Extra procedures or visits needed<br>E. Potential of harm for child<br>F. Privacy concerns for child<br>G. Overall interest in research topic was low | No, due to question format and lack of neutral framing. |
| **Q14** | When you have been approached to participate in a study including your child, how were you informed? | **Multiple Choice**<br>A. Our doctor notified us of a research study and asked if we wanted to participate<br>B. Flyer and information sheet in clinic<br>C. Contacted or telephoned by research coordinator or nurse<br>D. Someone told me about it who had participated<br>E. Not applicable, I've never been approached to participate in a study with my child | No, due to question format and lack of neutral framing. |
| **Q15** | In general, would you say that childhood research is an added benefit to society? | **Multiple Choice**<br>A. Yes<br>B. No<br>C. Don't know or unsure | No, due to question format and lack of neutral framing. |
| **Q16** | Do you agree or disagree with this statement: childhood research is an important tool to improving medical care? | **Multiple Choice**<br>A. Strongly agree<br>B. Agree<br>C. Neutral<br>D. Disagree<br>E. Strongly disagree | No, due to question format, including lack of both experience-focused responses and neutral framing. |
| **Q17** | Do you agree or disagree with this statement: childhood medical research is most beneficial to doctors rather than the general public? | **Multiple Choice**<br>A. Strongly agree<br>B. Agree<br>C. Neutral<br>D. Disagree<br>Strongly disagree | No, due to question format and lack of neutral framing. |
| **Q18** | Do you trust medical facilities with conducting childhood research? | **Multiple Choice**<br>A. Yes<br>B. No<br>C. Don't know | No, due to question format and lack of neutral framing. |
| **Q19** | What are some of your concerns about giving specimens? | **Multiple Choice**<br>A. Risk to child's health<br>B. specimens? Risk to child's privacy or security<br>C. Cost of participation<br>D. Compensation for participation<br>E. Lack of clear benefit to child's health<br>F. Lack of clear benefit to the community's health<br>G. Other | Yes, due to question format, concept clarity, experience focus, and neutral framing. |
| **Q20** | What do you feel is the barrier when conducting research on children? | **Multiple Choice**<br>A. Risk to child's health<br>B. specimens? Risk to child's privacy or security<br>C. Cost of participation<br>D. Compensation for participation<br>E. Lack of clear benefit to child's health<br>F. Lack of clear benefit to the community's health<br>G. Other | Yes, due to question format, concept clarity, experience focus, and neutral framing. |
| **Q21** | As applicable, how do you feel about answering this questionnaire for research studies? | **Free Response** | Yes, due to question format, concept clarity, experience focus, and neutral framing. |
| **Q22** | What suggestions do you have for research investigators to improve patient's and their family's experiences of enrolling in a study? | **Free Response** | Yes, due to question format, concept clarity, experience focus, and neutral framing. |

**Table S.4.** Hyperparameter value inputs for Hybrid Human-AI model for both UVA and TU. Values utilized to generate cluster labeling based on single-sentence theme clustering using GPT-4 omni model [2].

| Hyperparameter | Value |
| --- | --- |
| Temperature | 0.2 |
| Max_tokens | 4096 |
| Top_p | 1.0 |
| Frequency_penalty | 0.3 |
| Presence_penalty | 0.2 |

**Table S.5.**
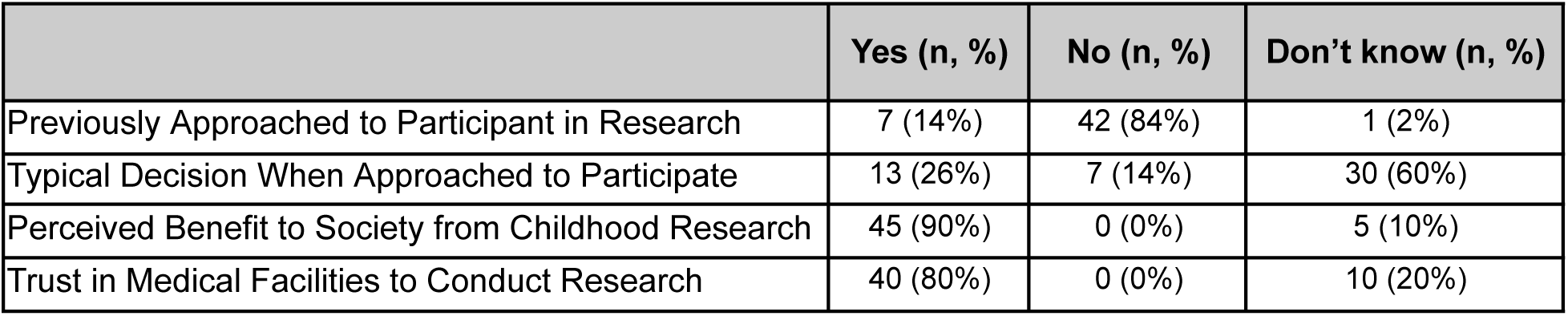
Caregiver perception about child participation in research at Tulane University.

|  | Yes (n, %) | No (n, %) | Don't know (n, %) |
| --- | --- | --- | --- |
| Previously Approached to Participant in Research | 7 (14%) | 42 (84%) | 1 (2%) |
| Typical Decision When Approached to Participate | 13 (26%) | 7 (14%) | 30 (60%) |
| Perceived Benefit to Society from Childhood Research | 45 (90%) | 0 (0%) | 5 (10%) |
| Trust in Medical Facilities to Conduct Research | 40 (80%) | 0 (0%) | 10 (20%) |

**Table S.6.**
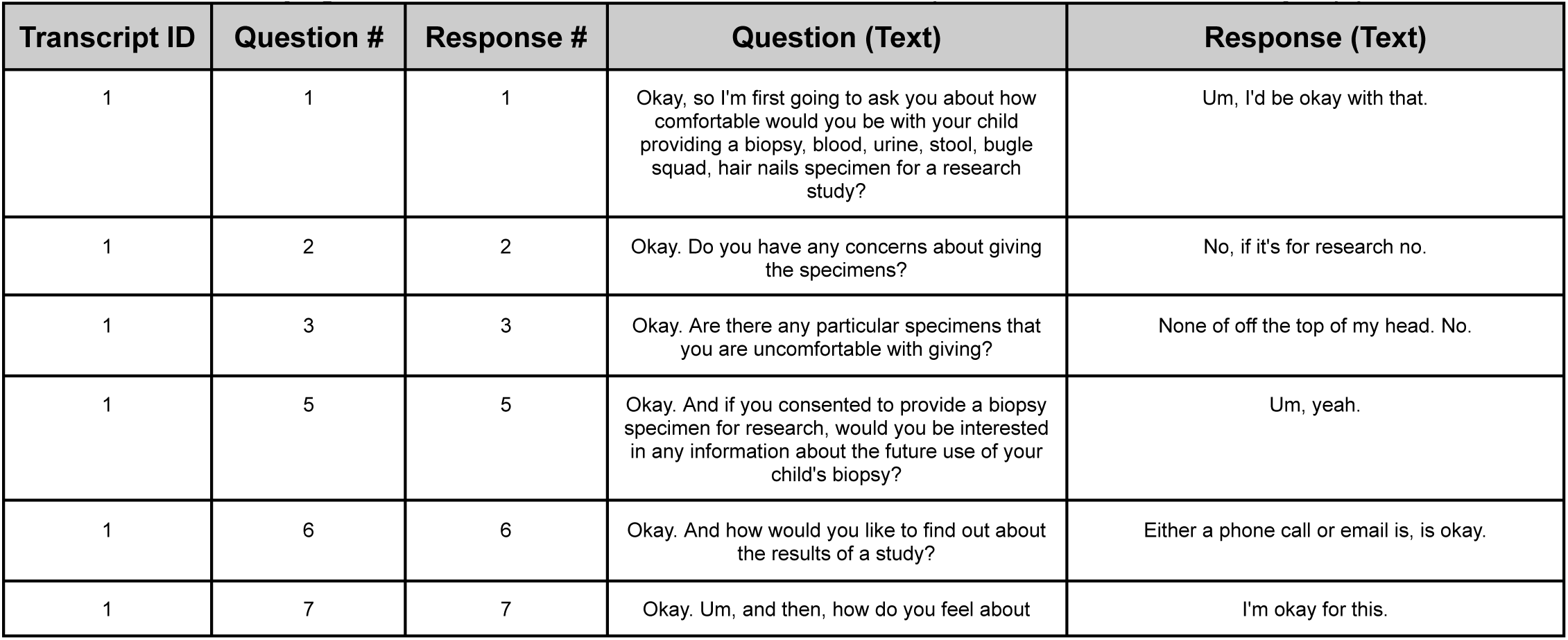
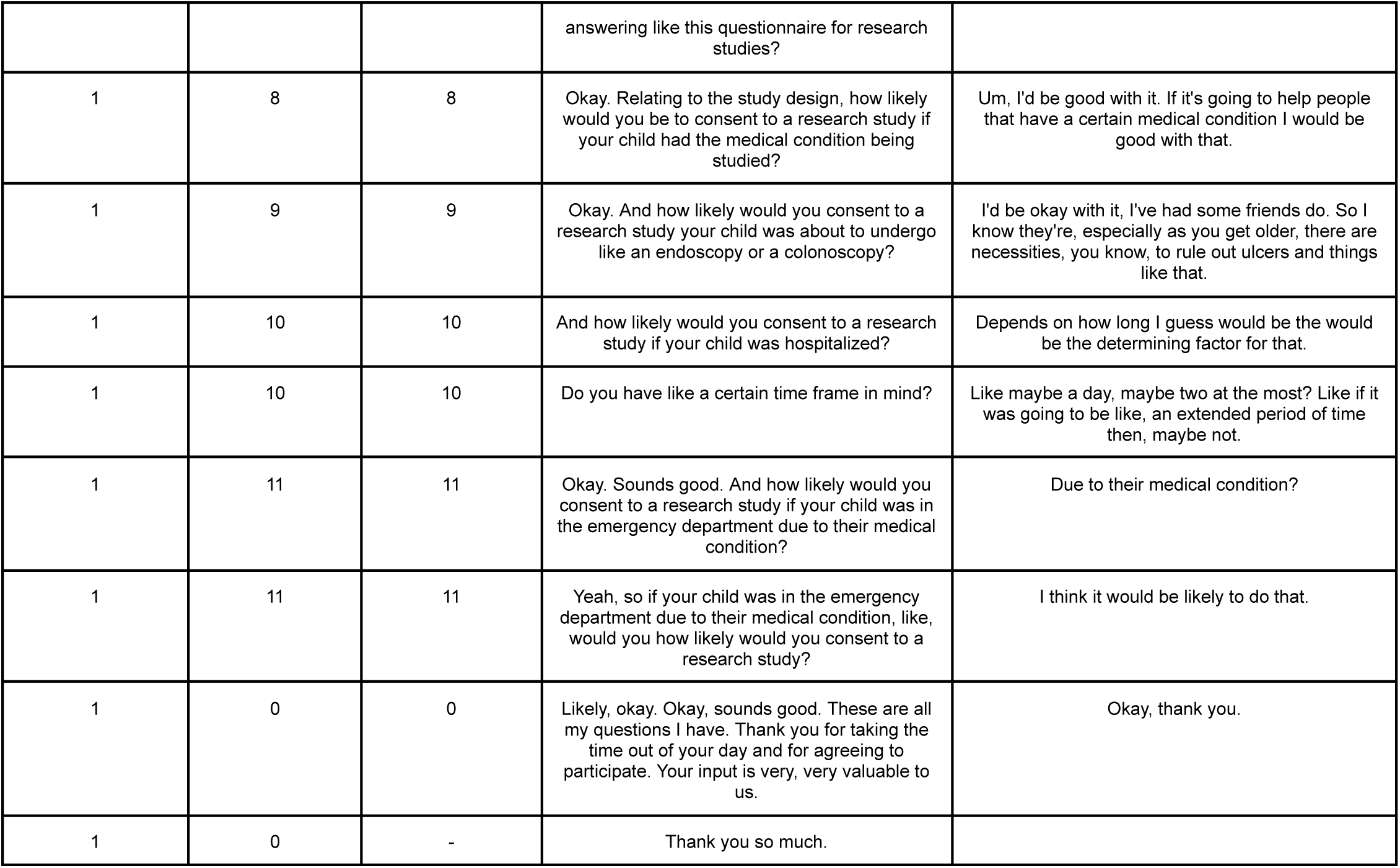
Example transcript from UVA Participant showing complete question–response pairs used in the analysis. This table displays a full example of the semi-structured interview dialogue, including all interviewer questions and corresponding participant responses for one guardian. These transcripts were used as input for GPT-4 [4]-based models to analyze consent likelihood, thematic content, emotional expression, and sentiment. Participant responses were used to assess underlying motivations, concerns, and emotional tone as part of the multi-model analytic pipeline.

**Table S.7.** Example transcript from TU Participant showing complete question–response pairs used in the descriptive-focused coding, which was used as an input for GPT-4 omni [10] for thematic analysis.

| Survey ID | Question # | Response # | Question (Text) | Response (Text) |
| --- | --- | --- | --- | --- |
| 1 | 19 | 19 | What are some of your concerns about giving the specimens? | Other |
| 1 | 20 | 20 | What do you feel is the barrier when conducting research on children? | Other |
| 1 | 21 | 21 | As applicable, how do you feel about answering this questionnaire for research studies? | Everyone should participate |
| 1 | 22 | 22 | What suggestions do you have for research investigators to improve patient and their family's experiences of enrolling in a study? | No |

**Table S.8:** List of descriptive-focused coding from TU codebook in NVivo using data extracted from surveys.

| **Descriptive-focused Code by Question** | **Number of respondents** |
| --- | --- |
| **Q1: What are some of your concerns about giving specimens?** | 50 |
| Financial or logistical burden of participation | 5 |
| Incentives and fair compensation | 2 |
| Potential harm to child's well-being | 15 |
| Privacy and data security concerns | 9 |
| Uncertain individual health benefits | 6 |
| Unclear community-level impact or relevance | 1 |
| Response: Other (no code) | 12 |
| **Q2: What do you feel is the barrier when conducting research on children?** | 50 |
| Practical and logistical barriers | 15 |
| Research value and justification | 3 |
| Safety and protection concerns | 25 |
| Response: Other (no code) | 7 |
| **Q3: As applicable, how do you feel about answering this questionnaire for research studies?** | 32 |
| Efficient survey | 1 |
| Inclusive participation | 1 |
| Parent and caregiver impressions | 2 |
| Perceived value to participants | 5 |
| Positive survey experience | 21 |
| Response: N/A or no actual response of feelings (no code) | 2 |
| **Q4: What suggestions do you have for research investigators to improve patient and their family's experiences of enrolling in a study?** |  |
| Diversify recruitment modalities | 1 |
| Financial compensation | 2 |
| Full disclosure of benefits and risks | 4 |
| Historical knowledge | 1 |
| Increase in clinic visit time | 1 |
| Inform parents about research opportunities specific to their child | 1 |
| Minimize risk to child's health | 3 |
| Scheduling | 1 |
| Study design | 1 |
| Transparent consent process | 2 |
| Response: None, N/A, or No (No code) | 12 |
| Response: Don't know or Not sure (No code) | 2 |

**Table S.9:** List of descriptive-focused coding from the UVA codebook in NVivo using data extracted from interviews transcribed using OtterAi.

| **Descriptive-focused Code by Question** | **Number of respondents** |
| --- | --- |
| **Q1 (combined questions 1 and 2)** | 42 |
| Amenable to child's participation | 33 |
| Concern regarding additional testing | 2 |
| Concern regarding invasive sample collection | 5 |
| Concern regarding privacy | 4 |
| Concern regarding risk of infection | 1 |
| Concern regarding risk of pain or discomfort | 1 |
| Concern regarding risk to child's health | 2 |
| Familiar with process of sample collection | 1 |
| Initial concerns regarding child's participation | 20 |
| Not familiar with process of sample collection | 1 |
| **Q2 (combined questions 3 and 4)** | 40 |
| Concern regarding sample collection | 18 |
| Risk of anesthesia | 3 |
| Risk of pain | 4 |
| Restriction on participation based on specimen type | 28 |
| **Q3 (combined questions 5 and 6)** | 42 |
| Interested in future use of specimen | 42 |
| Interested in results of current research study | 42 |
| **Q4** | 37 |
| Amenable to questionnaire completion | 11 |
| No concerns with type of questions asked | 26 |
| **Q5** | 42 |
| Amenable if child has condition being studied | 3 |
| Amenable to child participation in research of child's medical condition | 35 |
| Consent for Study Participation | 2 |
| Depends on child's willingness to participate | 1 |
| Not amenable to participate | 1 |
| **Q6** | 41 |
| Amenable if specimen collected during pre-planned procedure | 7 |
| Amenable to child's participation for upper endoscopy or colonoscopy for data collection | 31 |
| Consent depends on child's age | 1 |
| Consent depends on child's response | 1 |
| Not amenable to participate | 2 |
| **Q7** | 40 |
| Amenable to child participation in research study if hospitalized if admitted for at least 1 day. | 31 |
| Consent depends on invasiveness of specimen collection | 0 |
| Consent depends on reason for hospitalization | 6 |
| Consent depends on severity of child's condition | 3 |
| Not amenable to participate | 1 |
| **Q8** | 41 |
| Amenable to child's participation if in the ED due to medical conditions being studied. | 29 |
| Depends on child's willingness to participate | 1 |
| Ease of sample collection | 2 |
| Not amenable to participate | 1 |
| Reason for ER visit | 3 |
| Severity of medical condition | 3 |
| Time in the ER | 2 |
| Type of medical condition | 3 |

**Table S.10:**
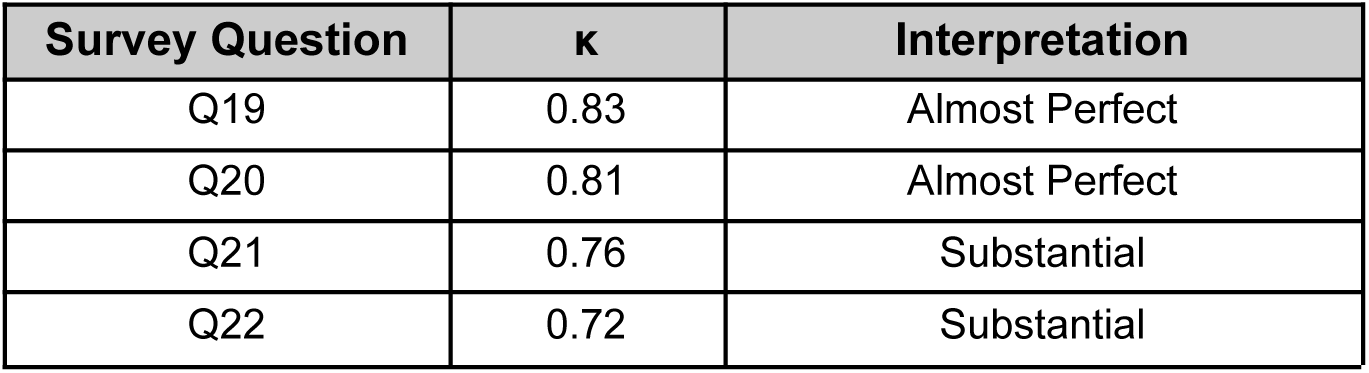
Cohen’s kappa agreement between GPT-coded (Fully Automated-AI approach) and human NVivo-coded (Hybrid Human AI approach) survey responses for TU participants across four thematic questions.

**Table S.11:** Cohen’s kappa agreement between GPT-coded (Fully Automated-AI approach) and human NVivo-coded (Hybrid Human-AI approach) interview responses for UVA participants across eight thematic questions.

| Interview Question | $\kappa$ | Interpretation |
| --- | --- | --- |
| Q1 | 0.79 | Substantial |
| Q2 | 0.77 | Substantial |
| Q3 | 0.84 | Almost Perfect |
| Q4 | 0.91 | Almost Perfect |
| Q5 | 0.74 | Substantial |
| Q6 | 0.72 | Substantial |
| Q7 | 0.70 | Substantial |
| Q8 | 0.69 | Substantial |

---

**Figure S.1.**
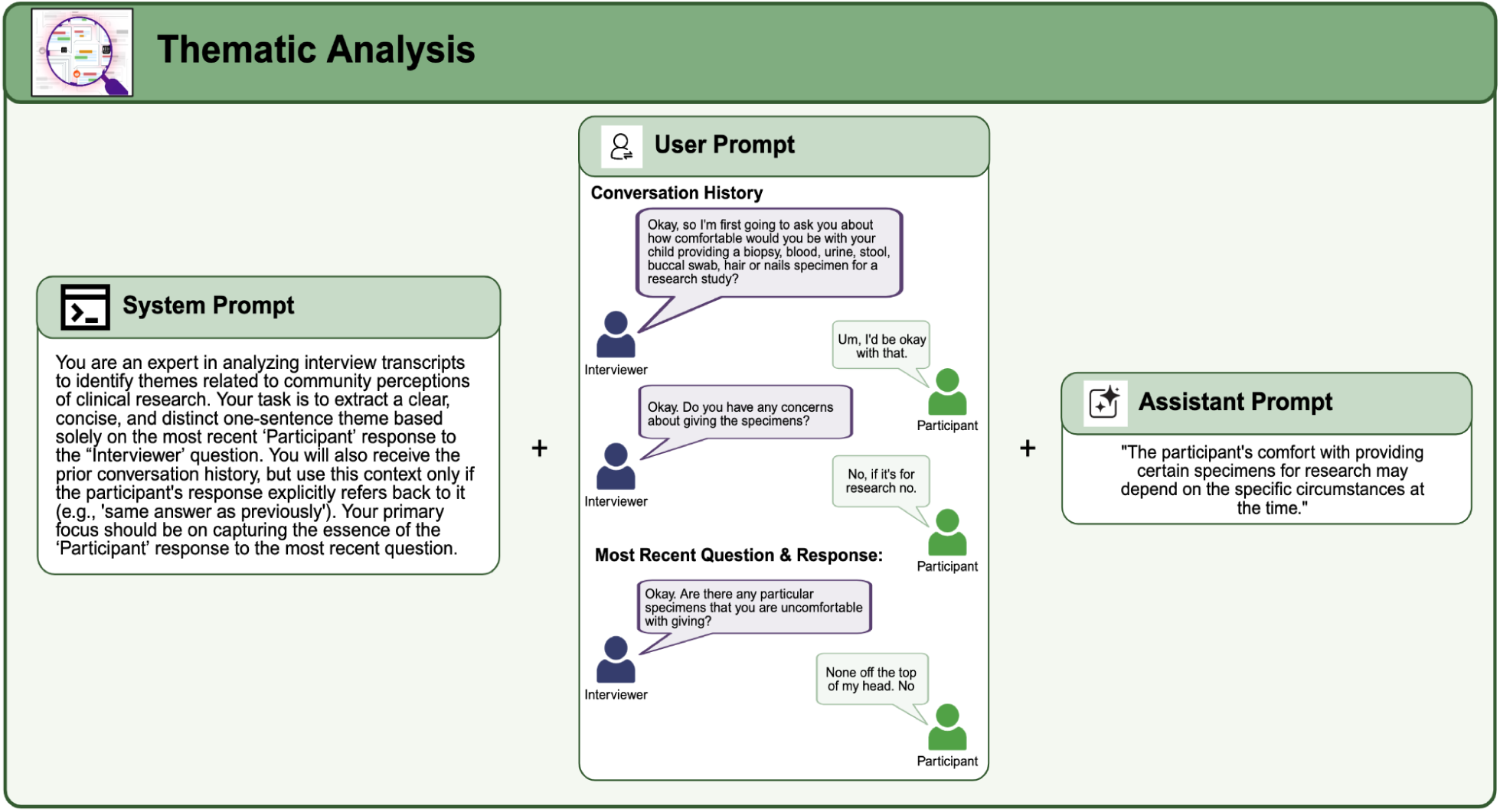
Overview of the prompting (**A-**system, **B-**user, **C-**assistant) framework used for GPT-4 [4]-based thematic analysis to generate a single-sentence theme for each question-response pair. The **System Prompt (left panel)** was adapted to instruct the model to generate concise thematic summaries. **Panel B-** The **User Prompt** provides the full conversation context and highlights the most recent interview question and participant answer. **Panel C-** The **Assistant prompt** represents the model’s expected output format—a single, coherent thematic sentence capturing the essence of the participant’s latest response.

**Figure S.2.**
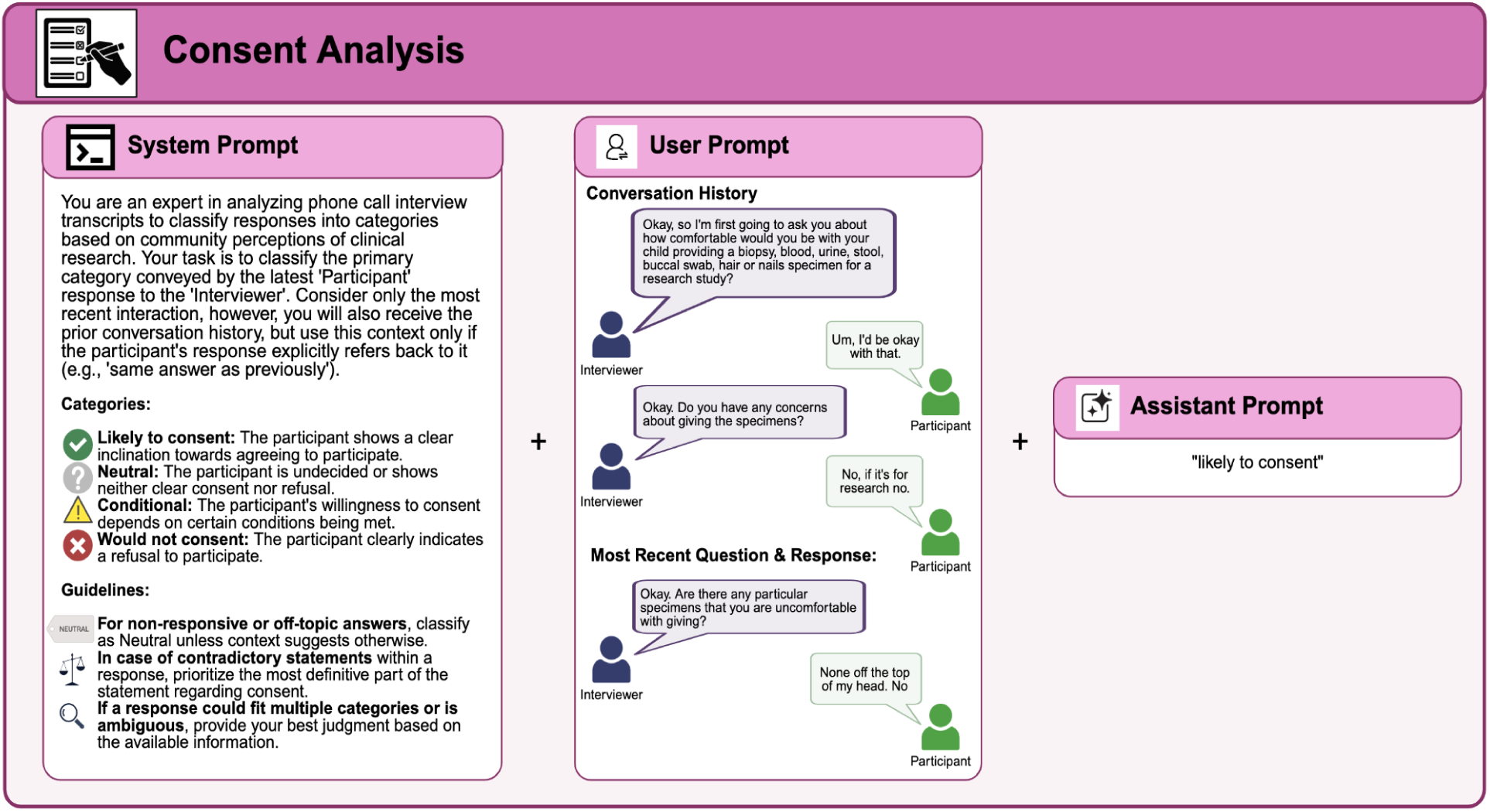
Overview of the prompting (**A-**system, **B-**user, **C-**assistant) framework used for GPT-4 [4]-based consent classification to generate predefined consent categories per question. **Panel A-** The **System Prompt** establishes GPT-4’s [4] role as an “expert/AI” tasked with assigning one of four labels—Likely to consent, Neutral, Conditional, or Would not consent—based on the participant’s most recent statement, while also providing guidance for handling ambiguity, contradictions, or non-responses; **Panel B-** The **User Prompt** provides the model with the conversation history and highlights the most recent interviewer question and participant response to be evaluated; **Panel C-** The **Assistant prompt** represented the model’s expected output format, it was a single consent classification label, such as “likely to consent.”

**Figure S.3.**
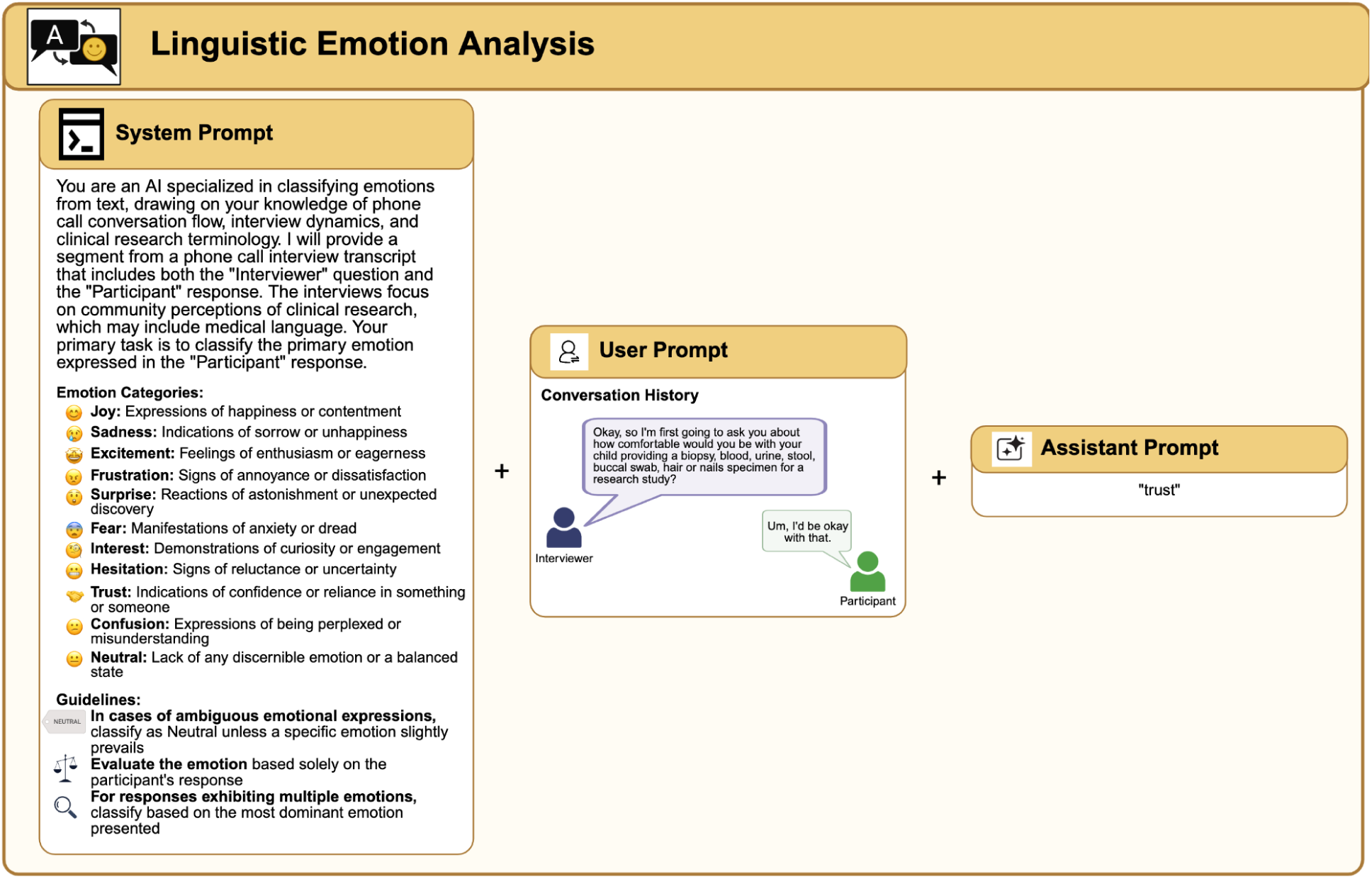
Overview of the prompting (**A-**system, **B-**user, **C-**assistant) framework used for GPT-4 [4]-based linguistic emotion classification to classify the primary emotion expressed by participants in phone call interviews about community perceptions of clinical research. **Panel A-** The **System Prompt** instructs GPT-4 [4] to act as an emotion classification expert, focusing on identifying a single dominant emotion based on the participant’s most recent response. It provides a list of emotion categories—including joy, trust, sadness, fear, and others—as well as guidelines for resolving ambiguous or multi-emotion responses. **Panel B-** The **User Prompt** presents a transcript segment containing the interviewer’s question and the participant’s response. **Panel C-** The **Assistant prompt** shows GPT-4’s [4] output: a single-word label identifying the most salient emotion conveyed in the participant’s statement—in this example, “trust.”

**Figure S.4.**
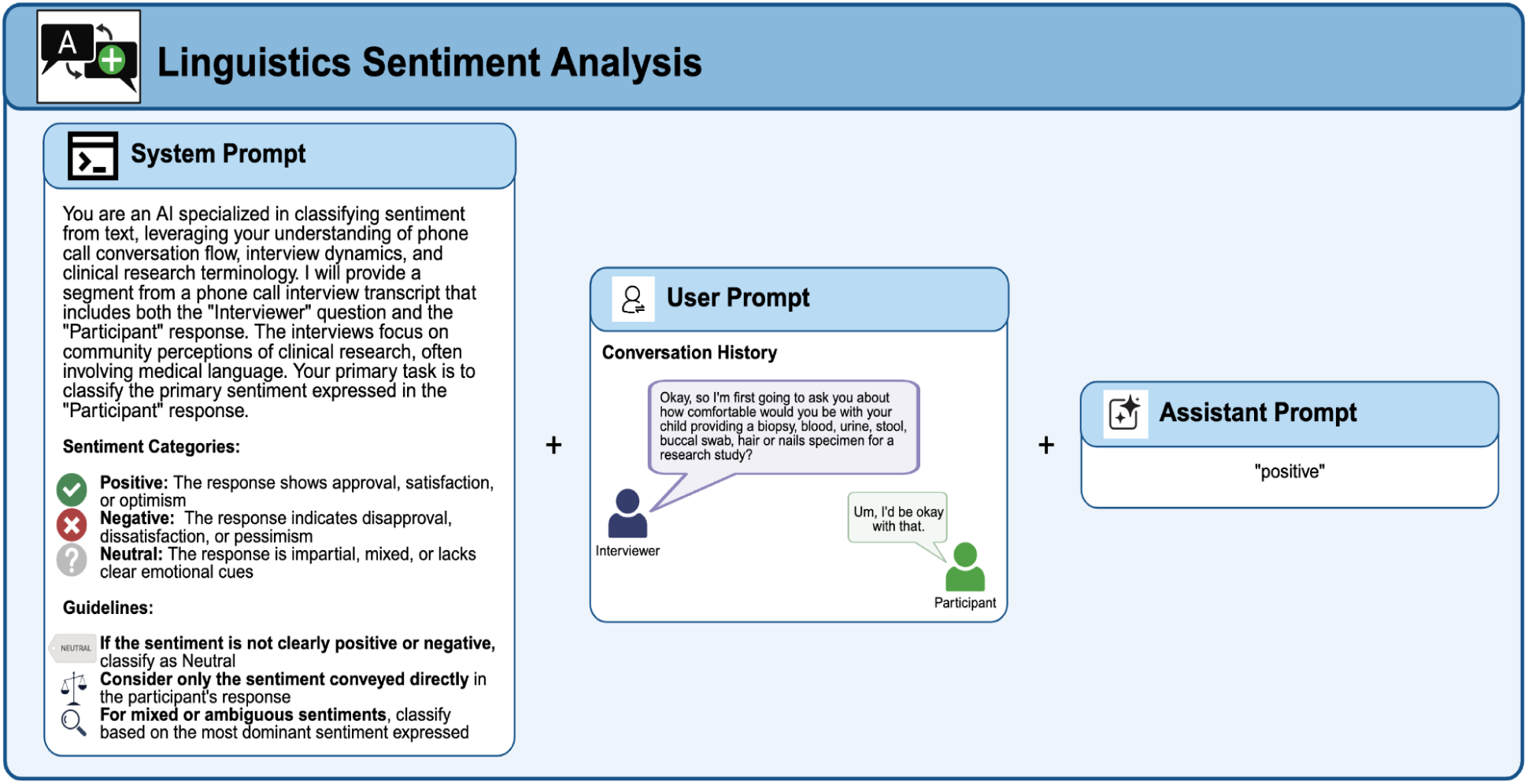
Overview of the prompting (**A-**system, **B-**user, **C-**assistant) framework used for GPT-4 [4]-based sentiment analysis to classify sentiment in participant responses during phone call interviews about clinical research. **Panel A-** The **System Prompt** defines GPT-4’s [4] task: to identify whether a participant’s most recent response conveys a positive, negative, or neutral sentiment based on emotional tone, approval, or disapproval. It also provides guidance for handling mixed or ambiguous sentiments. **Panel B-** The **User Prompt** includes the interview question and the participant’s response. **Panel C-** The **Assistant prompt** shows GPT-4’s [4] output—a single sentiment label, such as “positive,” inferred from the participant’s reply.

**Figure S.5.**
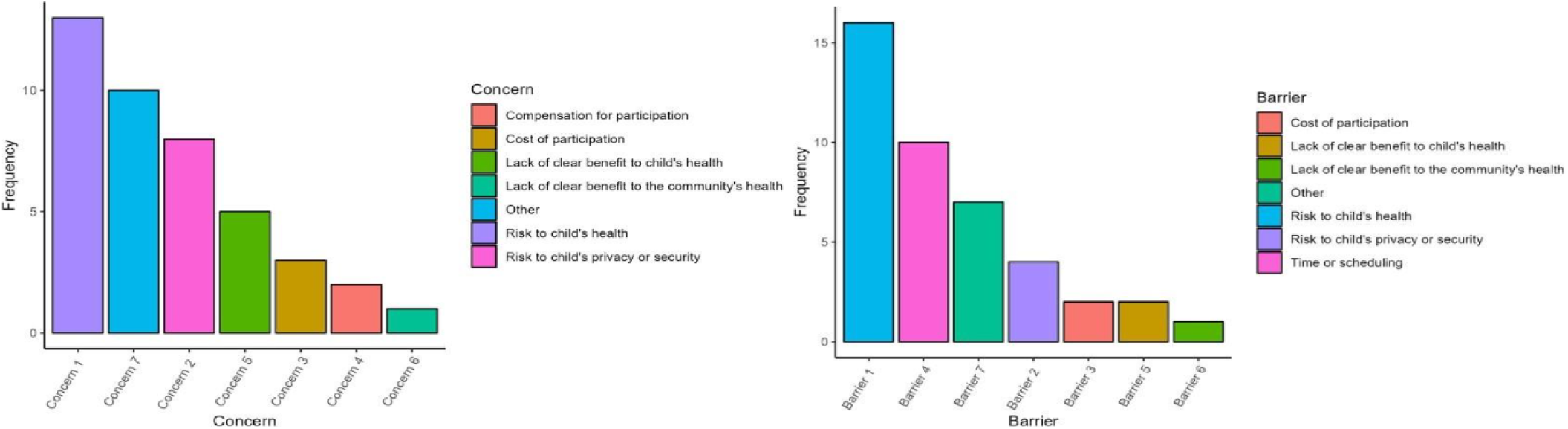
**A)** Caregivers perceived concerns about child participation in research at TU. **B)** Caregivers perceived barriers about child participation in research at TU.

**Figure S.6.**
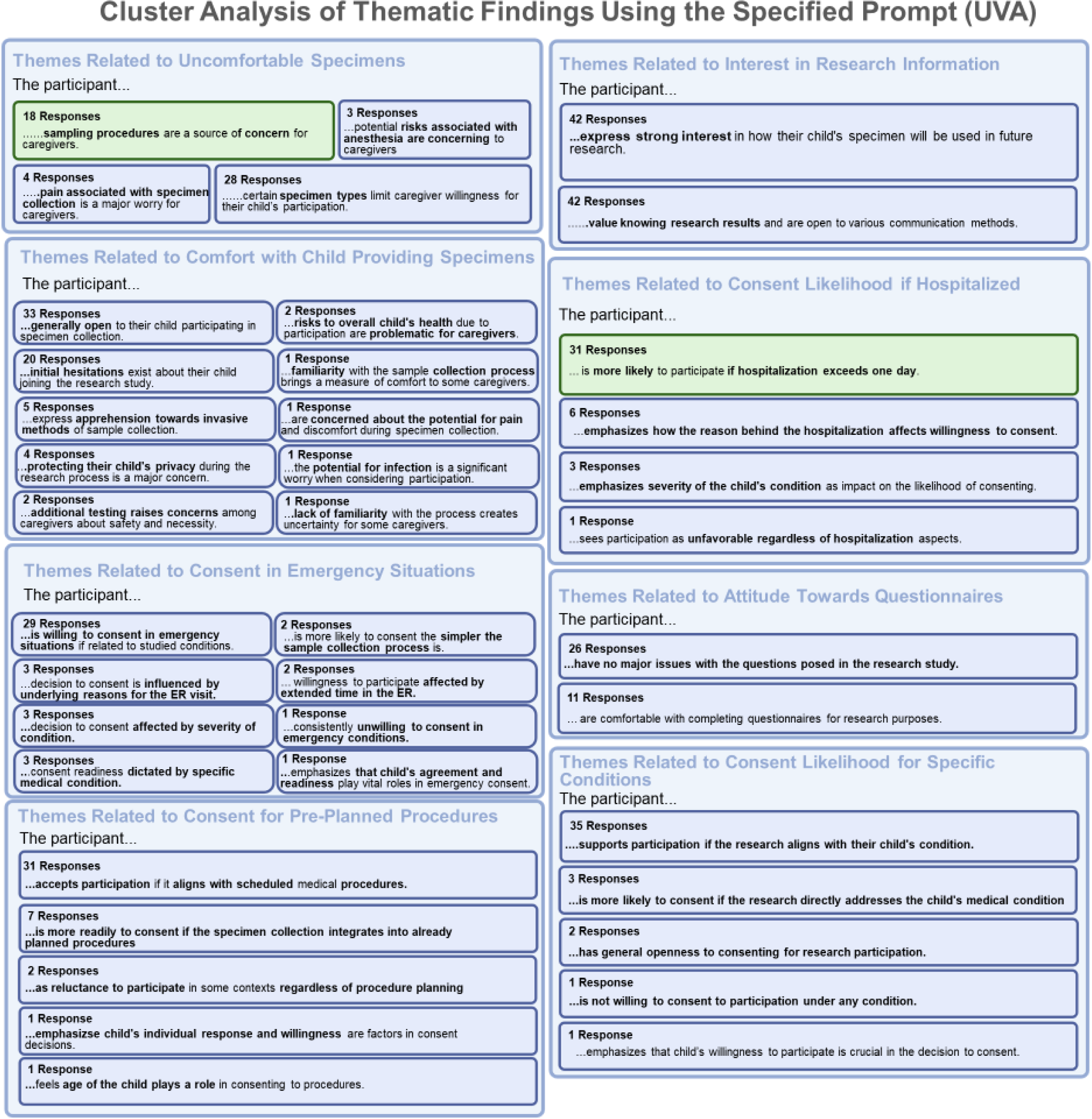
Themes identified from cluster analysis of UVA transcripts using a hybrid human-AI model with a site-specific prompt.

**Figure S.7.**
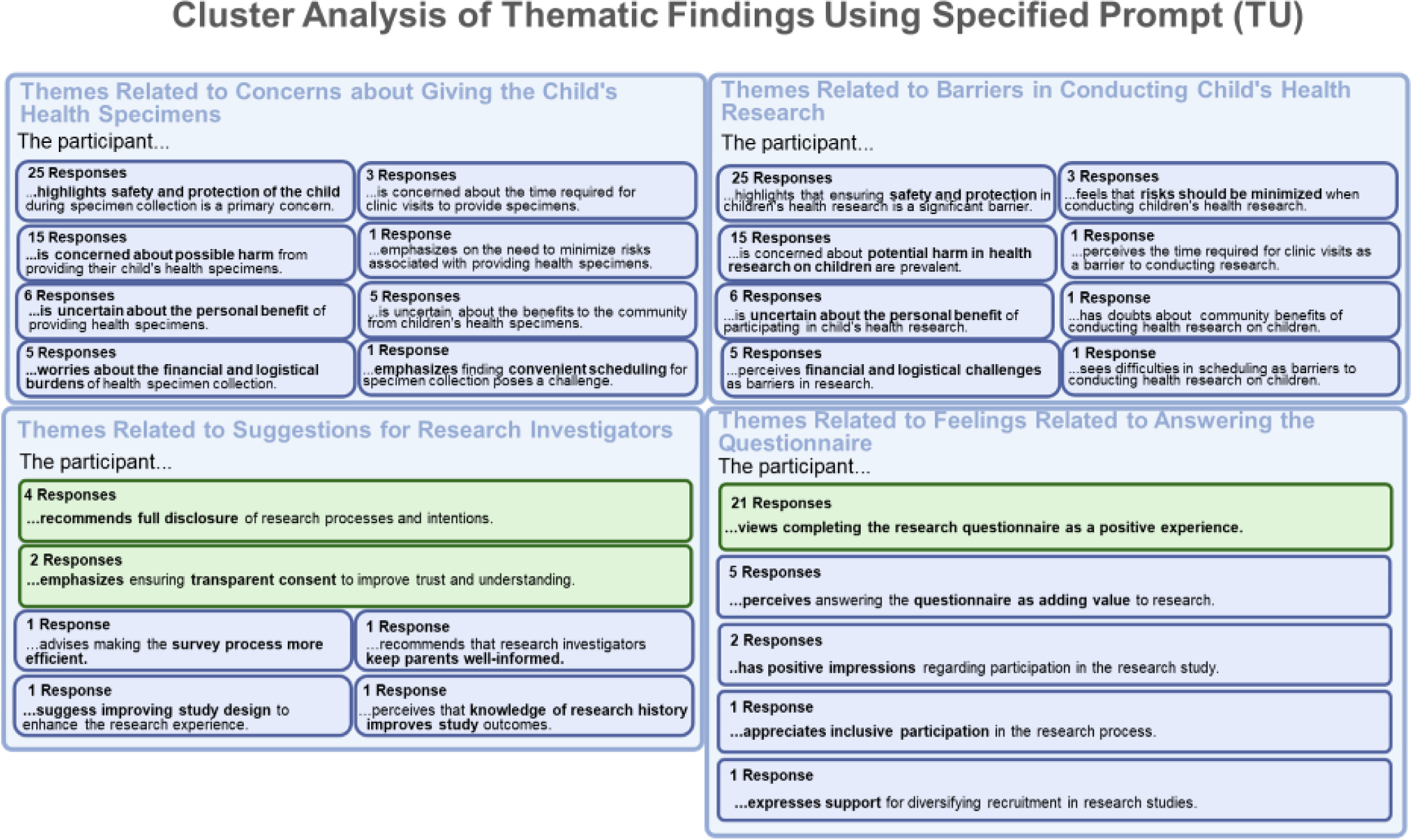
Themes identified from cluster analysis of TU question-response pairs using a hybrid human-AI model with a site-specific prompt.

**Figure S.8.**
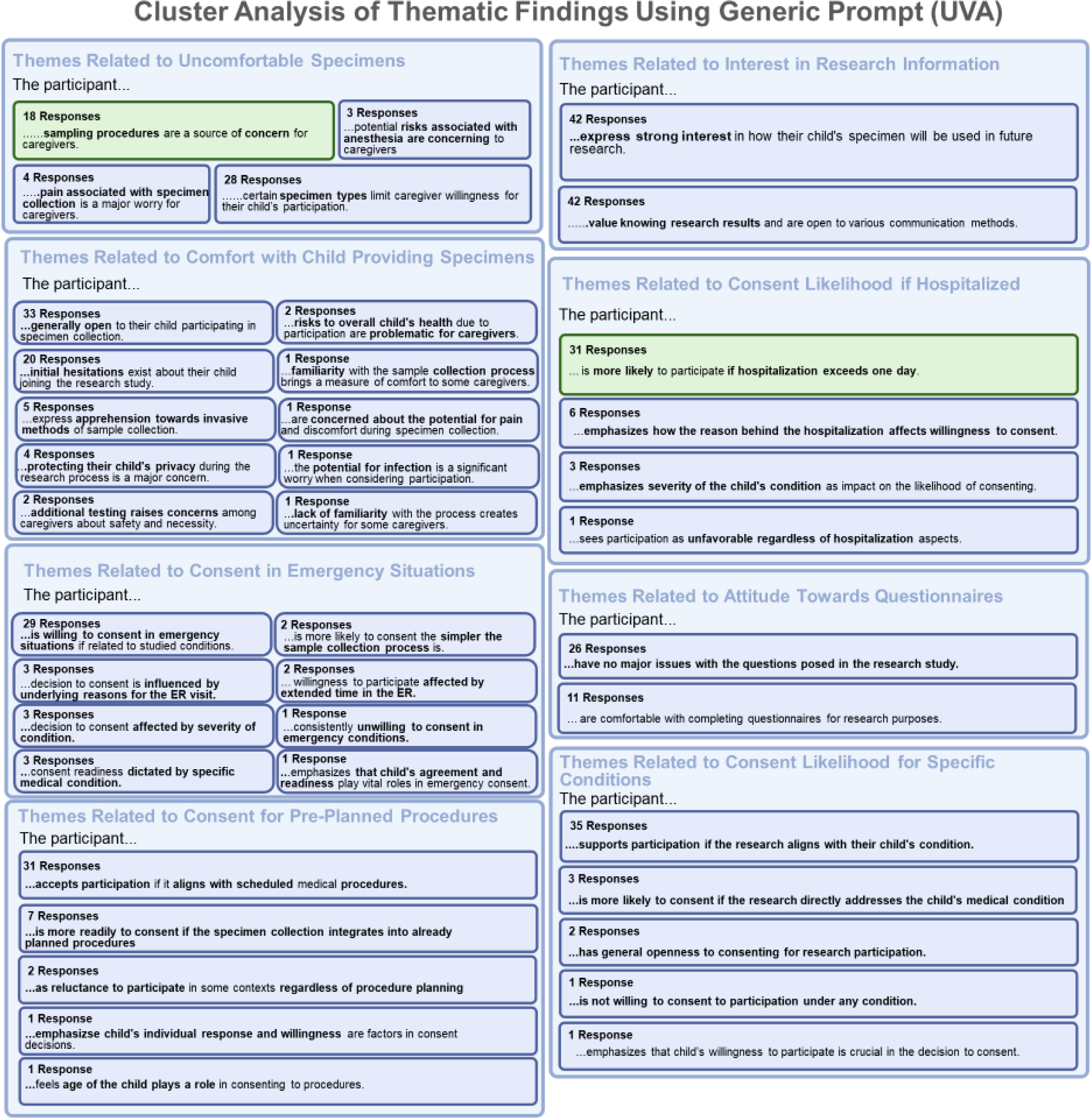
Themes identified from cluster analysis of UVA transcripts using a hybrid human-AI model with a generic prompt.

**Figure S.9.**
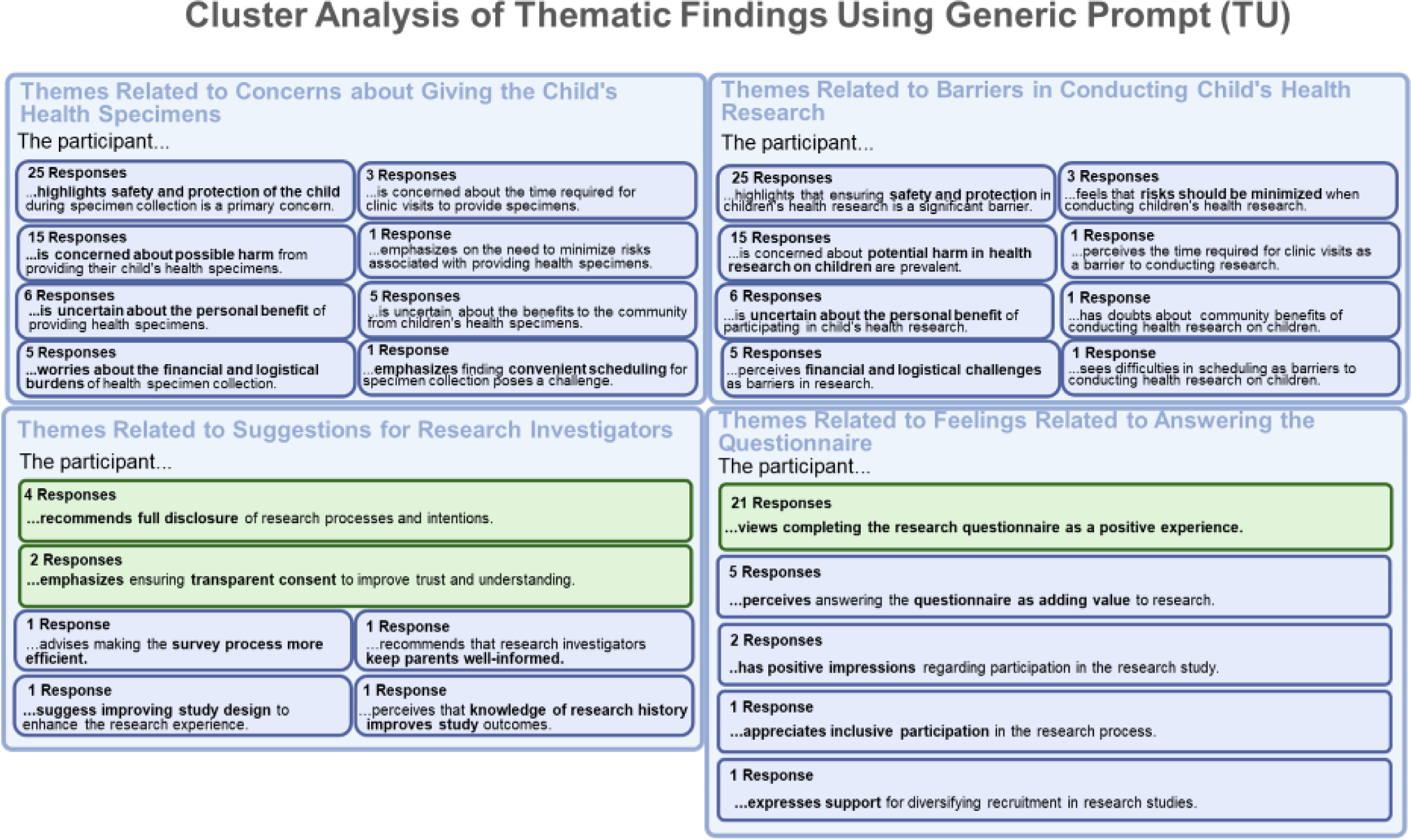
Themes identified from cluster analysis of TU question-response pairs using a Hybrid-Human AI model with a generic prompt.

**Figure S.10.**
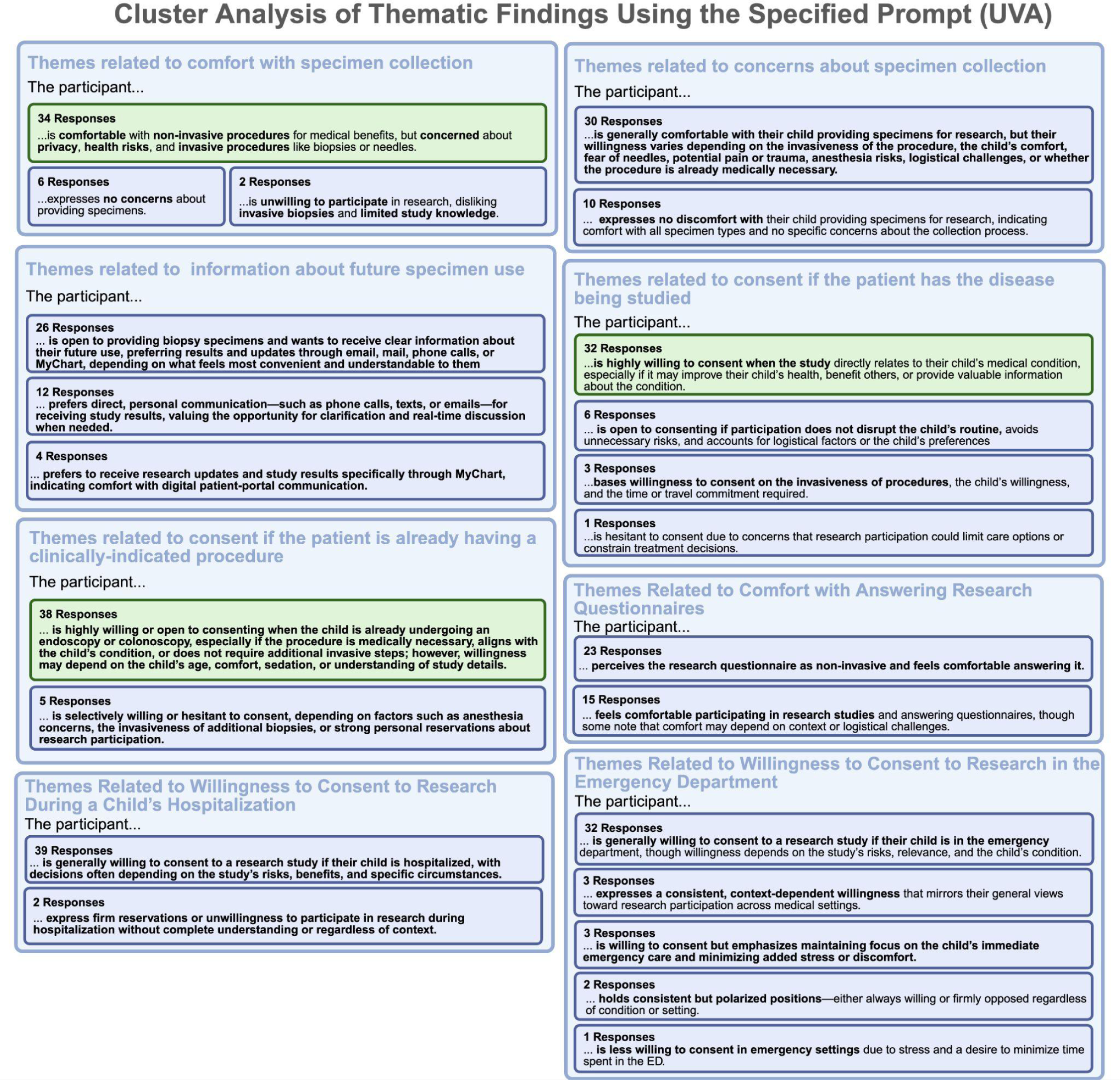
Themes identified from cluster analysis of UVA transcripts using an automated-AI model with a site-specific prompt.

**Figure S.11.**
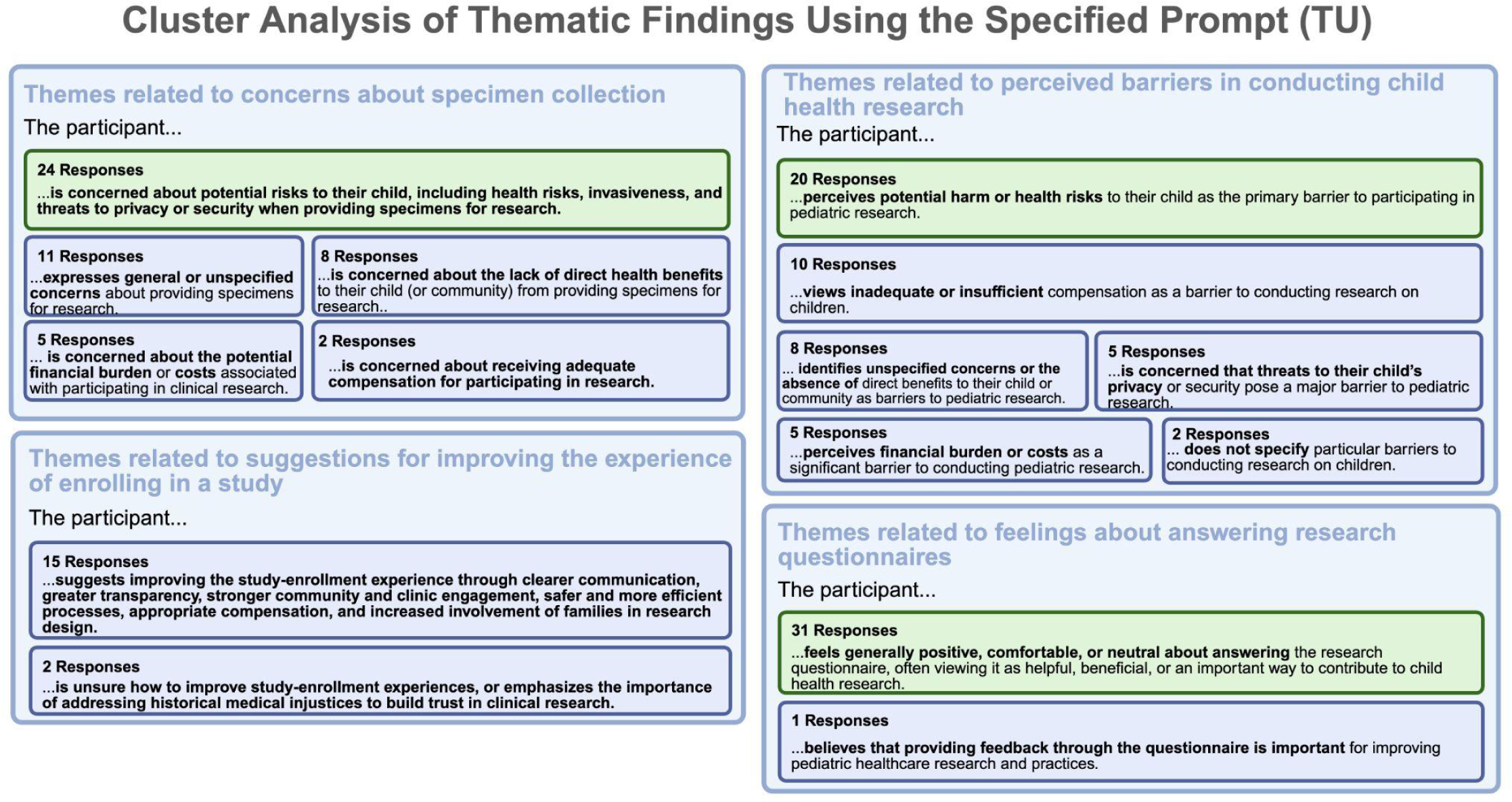
Themes identified from cluster analysis of TU question-response pairs using an automated-AI model with a site-specific prompt.

**Figure S.12.**
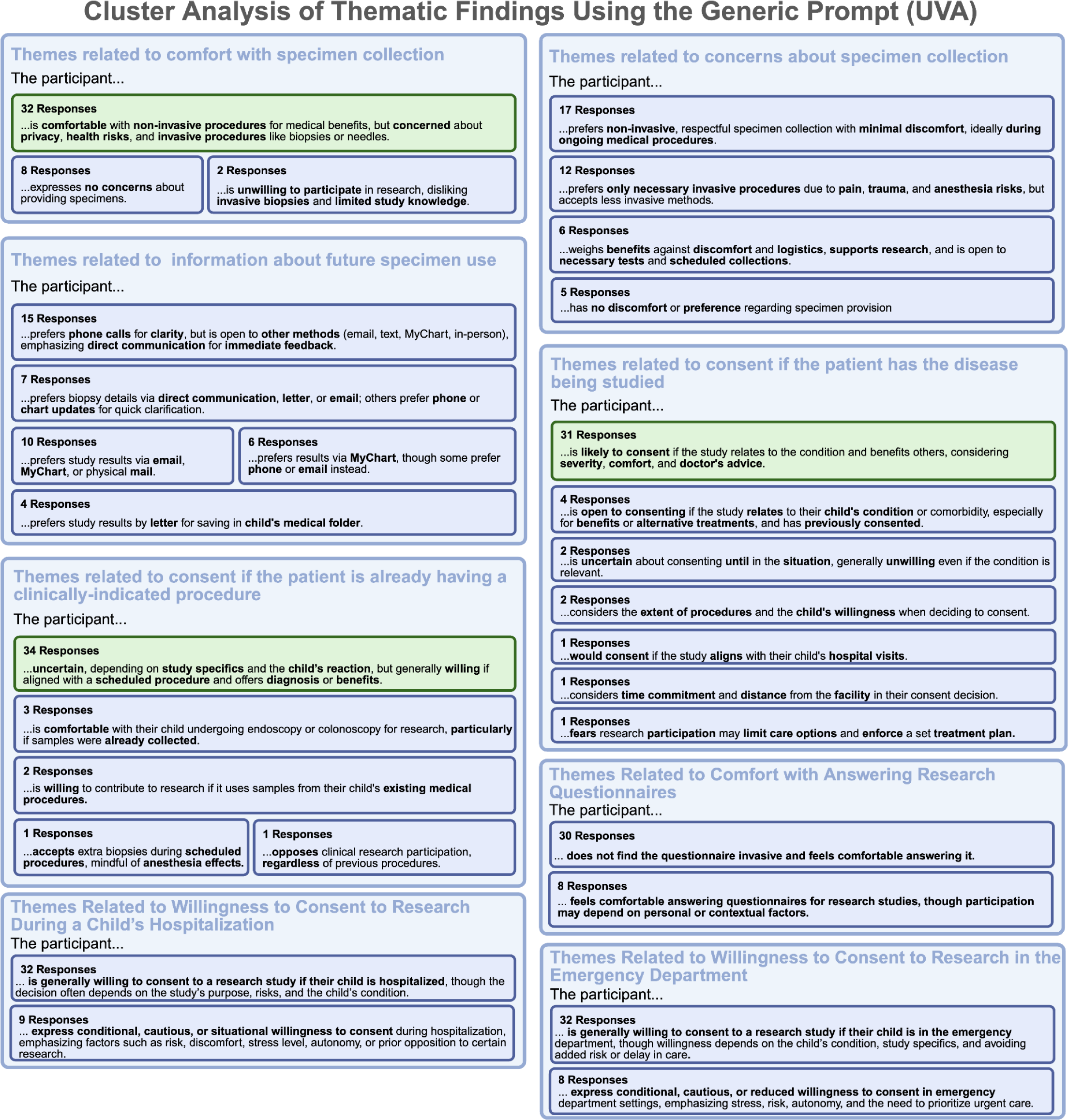
Themes identified from cluster analysis of UVA transcripts using an automated-AI model with a generic prompt.

**Figure S.13.**
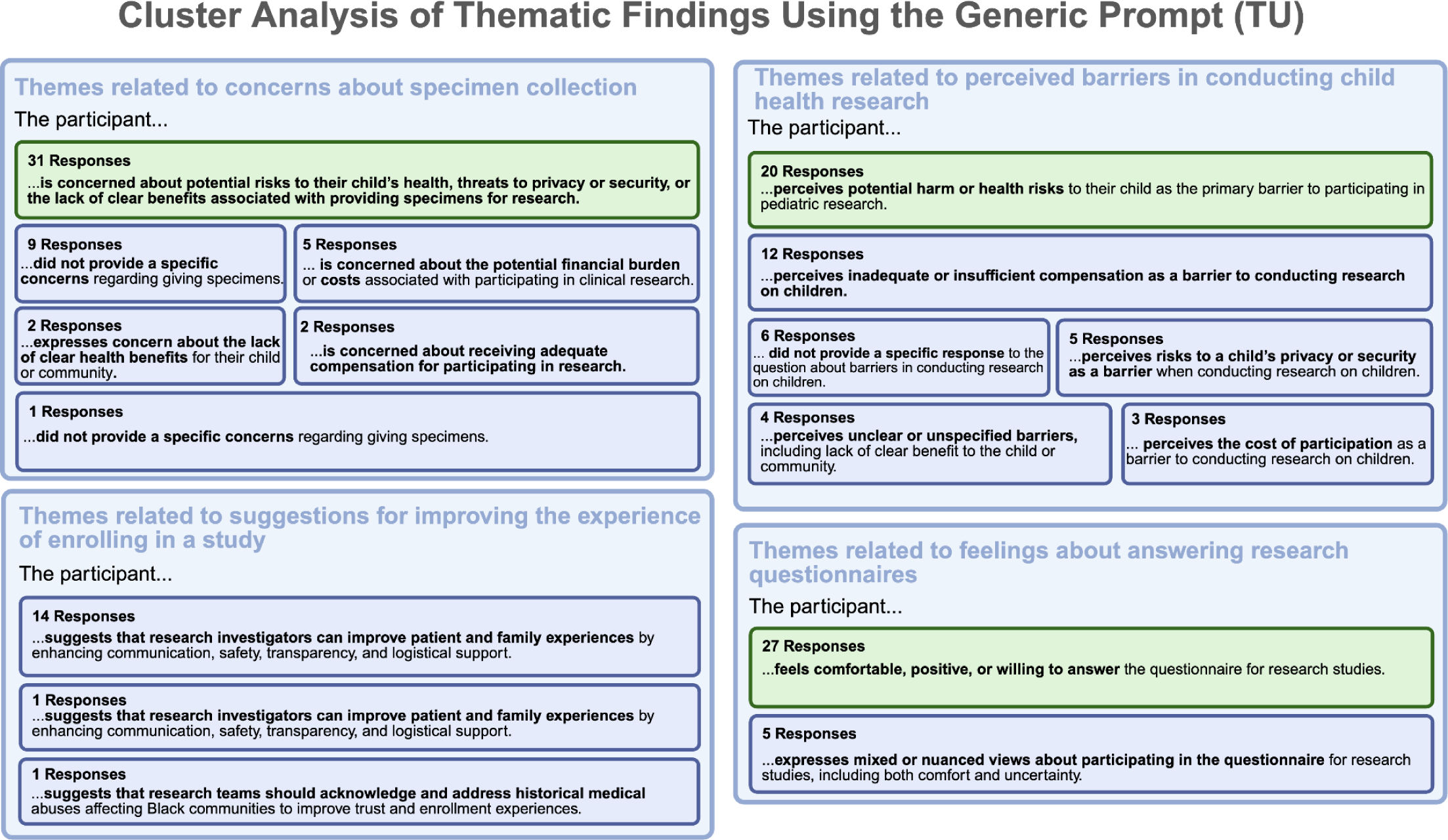
Themes identified from cluster analysis of TU question-response pairs using an automated-AI model with a generic prompt.

**Figure S.14.**
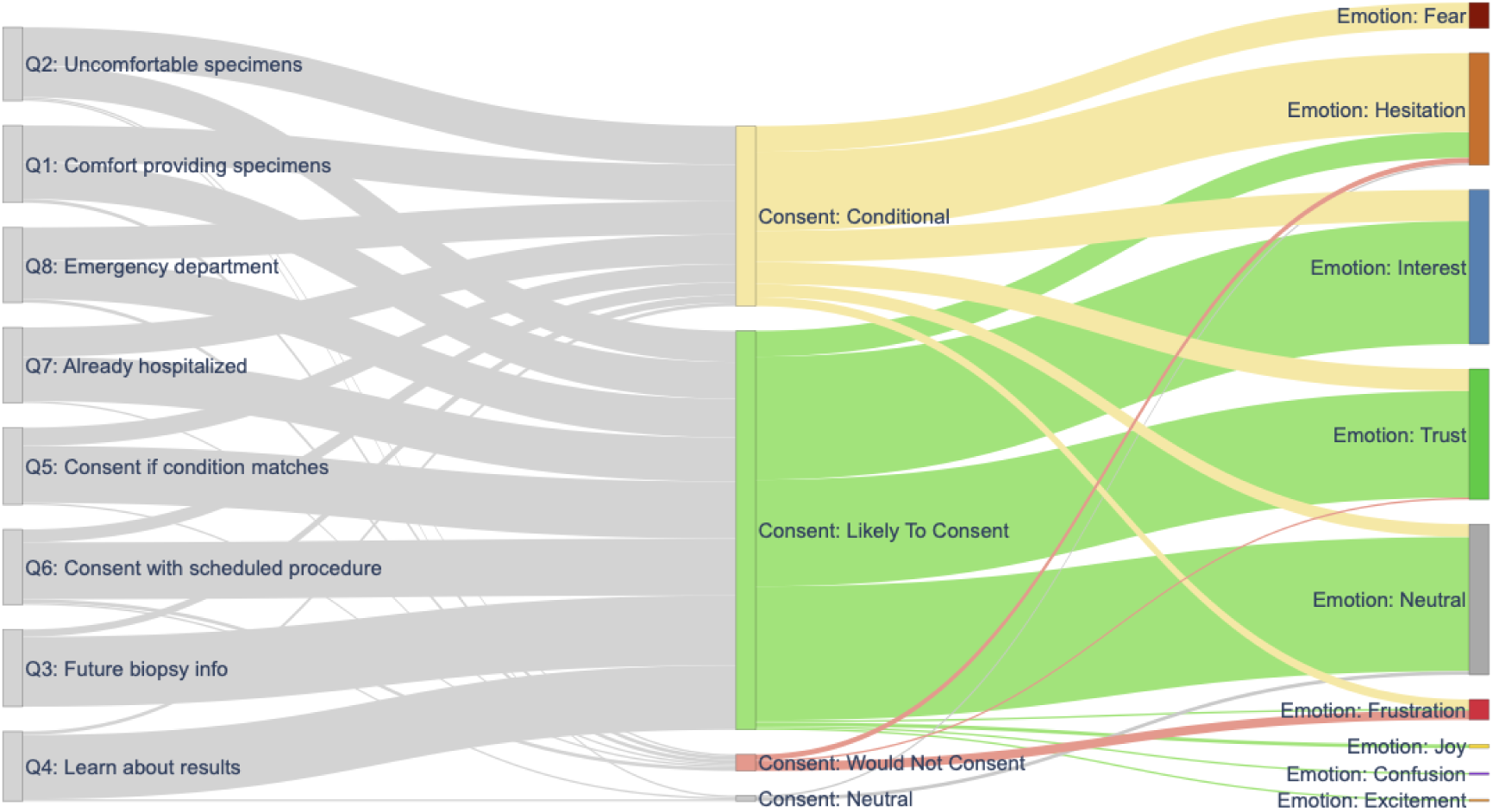
Flow of consent categories and dominant emotions across grouped interview questions. This Sankey diagram visualizes the relationship between consent classification and emotional expression in participants’ responses, as determined by GPT-4–based linguistic emotion analysis. Each band represents the number of responses transitioning from interview question groups (Q1–Q8) to consent categories—Likely to consent, Conditional, Neutral, and Would not consent—and then to dominant emotions. “Likely to consent” responses were most frequently associated with trust and interest, while conditional responses showed more hesitation and occasional fear or confusion, reflecting uncertainty toward participation. Negative emotional tones such as frustration and fear were rare, indicating overall confidence and receptivity among participants toward research engagement.

**Supplementary Figure S.15.**
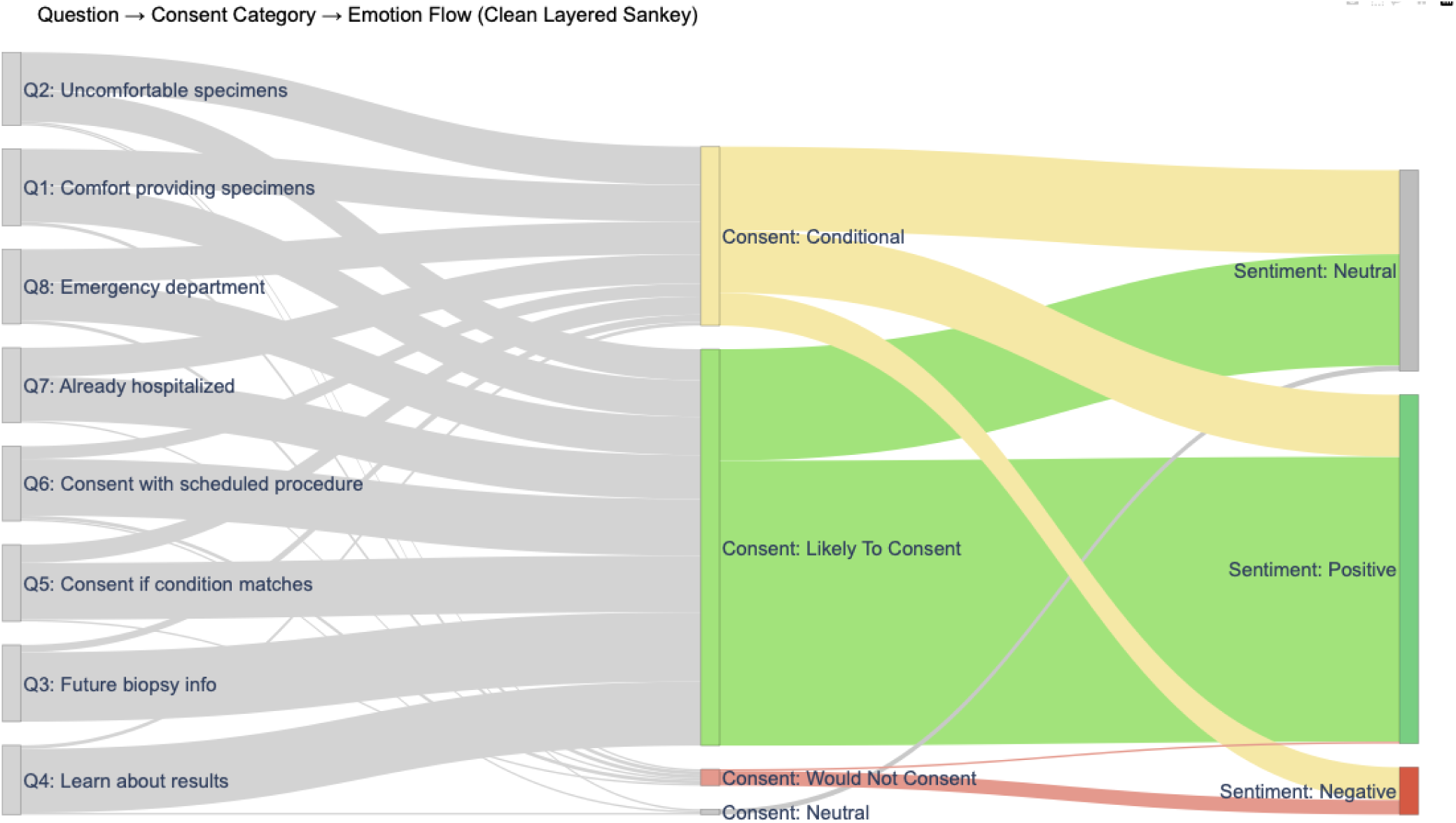
Integrated visualization of consent categories and sentiment distribution across interview questions. This Sankey diagram illustrates the flow from grouped interview questions (Q1–Q8) to consent classifications and corresponding sentiment outcomes, as determined by GPT-4–based automated models. The width of each band represents the relative number of responses. Most responses across questions mapped to the “Likely to consent” category, which was predominantly associated with positive sentiment (green). Conditional responses showed a more balanced distribution between neutral and positive sentiment (yellow), while “Would not consent” responses were infrequent and primarily aligned with negative sentiment (red).

## Notes

### Competing Interest Statement

The authors have declared no competing interest.

### Author Declarations

Institutional Review Board of the University of Virginia gave ethical approval for this work. Institutional Review Board of Tulane University gave ethical approval for this work.

## REFERENCES

1. Hare, M. M., Walker, C. S., Sarver, D. E., Lim, C. S., Brown, D. C., & Annett, R. D. (2023). Assessing attitudes towards pediatric research participation across diverse populations: Psychometric properties of a novel tool. Contemporary clinical trials, 133, 107321. 10.1016/j.cct.2023.107321

2. Slotkowski, R. A., Delair, S. F., & Neemann, K. A. (2022). Cross-sectional survey of parental barriers to participation in pediatric participant research registries. PloS one, 17(5), e0268553. 10.1371/journal.pone.0268553

3. Li, X., Keown-Stoneman, C. D. G., Borkhoff, C. M., Wong, P. D., Arafeh, D., Tavares, E., Thadani, S., Maguire, J. L., Birken, C. S., & TARGet Kids! Collaboration (2023). Factors associated with research participation in a large primary care practice-based pediatric cohort: Results from the TARGet Kids! longitudinal cohort study. PloS one, 18(4), e0284192. 10.1371/journal.pone.0284192

4. Kendel, N. E., Belsky, J. A., Stanek, J. R., Streby, K. A., & Shah, N. (2022). Caregiver Perspectives on Patient Participation in Biological Pediatric Cancer Research. Children (Basel, Switzerland), 9(6), 901. 10.3390/children9060901

5. Shanwetter Levit, N. and Saban, M., 2025. When investigator meets large language models: a qualitative analysis of cancer patient decision-making journeys. NPJ digital medicine, 8(1), p.336.

6. Mathis WS, Zhao S, Pratt N, Weleff J, De Paoli S. Inductive thematic analysis of healthcare qualitative interviews using open-source large language models: How does it compare to traditional methods?. Comput Methods Programs Biomed. 2024;255:108356. doi:10.1016/j.cmpb.2024.108356

7. The Audacity Team. Audacity [computer software]. Version 3.5.0. Pittsburgh (PA): The Audacity Team; 2024 [cited 2024]. Available from: https://www.audacityteam.org

8. Otter.ai Inc. *Otter.ai* [Internet]. Los Altos (CA): Otter.ai Inc; [cited 2024]. Available from: https://otter.ai

9. Riaz A, Bhamani S, Ahmed S, Umrani F, Jakhro S, Qureshi AK, et al. Barriers and facilitators to exclusive breastfeeding in rural Pakistan: a qualitative exploratory study. Int Breastfeed J. 2022;17(1):59. DOI: 10.1186/s13006-022-00495-4.

10. OpenAI. *GPT*-*4* [computer program]. Model 0613. San Francisco (CA): OpenAI; 2023 [cited 2024]. Available from: https://platform.openai.com

11. Reimers N, Gurevych I. SentenceTransformer [computer program]. Darmstadt (DE): UKP Lab; [cited 2024]. Available from: https://www.sbert.net

12. Reimers N, Gurevych I. Sentence-BERT: Sentence embeddings using Siamese BERT-networks. arXiv:1908.10084 [Preprint]. 2019 Aug 27.

13. Devlin J, Chang M-W, Lee K, Toutanova K. BERT: Pre-training of deep bidirectional transformers for language understanding. arXiv:1810.04805 [Preprint]. 2018 Oct 11.

14. Sentence-transformers. *all*-*MiniLM*-*L12*-*v2* [computer program]. Paris (FR): Hugging Face Inc.; [cited 2024]. Available from: https://huggingface.co/sentence-transformers/all-MiniLM-L12-v2

15. IBM Corp. *Watson Natural Language Understanding API* [computer program]. Version 2021-08-01. Armonk (NY): IBM Corp.; 2021 [cited 2024]. Available from: https://cloud.ibm.com/docs/natural-language-understanding

16. Liu Y, Ott M, Goyal N, et al. RoBERTa: A robustly optimized BERT pretraining approach. arXiv:1907.11692 [Preprint]. 2019 Jul 26.

17. cardiffnlp. *twitter*-*roberta*-*base*-*sentiment* [computer program]. Paris (FR): Hugging Face Inc.; [cited 2024]. Available from: https://huggingface.co/cardiffnlp/twitter-roberta-base-sentiment

18. Lumivero. (2025). *NVivo (Version 15.3.1)* [Computer software]. Denver (CO): QRS International; [cited 2025]. Available from: https://lumivero.com/products/nvivo/

19. Landis, J. R., & Koch, G. G. (1977). The measurement of observer agreement for categorical data. Biometrics, 33(1), 159–174.

20. Miguel A. Pena, Melicia Y. Whitley, Anirudh Sudarshan, Patricia Ellen Grant, Ravi R. Thiagarajan, Efren J. Flores, Valerie L. Ward; Strategies to Increase Enrollment and Retention in Pediatric Clinical Research: A Scoping Review. Pediatrics September 2025; 156 (Supplement 1): e2025070739O. 10.1542/peds.2025-070739O

21. Kraft, S. A., Porter, K. M., Sullivan, T. R., Anderson, E. E., Garrison, N. A., Baker, L., Smith, J. M., & Weiss, E. M. (2022). Relationship building in pediatric research recruitment: Insights from qualitative interviews with research staff. Journal of clinical and translational science, 6(1), e138. 10.1017/cts.2022.469

22. Fehring, L., Frings, J., Rust, P., Kempny, C., Thürmann, P. A., & Meister, S. (2025). Extension of the Consolidated Criteria for Reporting Qualitative Research Guideline to Large Language Models (COREQ+LLM): Protocol for a Multiphase Study. JMIR research protocols, 14, e78682. 10.2196/78682

23. PA Harris, R Taylor, R Thielke, J Payne, N Gonzalez, JG. Conde, Research electronic data capture (REDCap) – A metadata-driven methodology and workflow process for providing translational research informatics support, J Biomed Inform. 2009 April; 42(2):377–81.

